# Weight Trajectories and Cancer Risk: A Pooled Cohort Study

**DOI:** 10.64898/2026.04.23.26351553

**Authors:** Anton Nilsson, Marisa da Silva, Huyen T. Le, Christel Häggström, Jens Wahlström, Karl Michaëlsson, Ylva Trolle Lagerros, Sven Sandin, Patrik K. E. Magnusson, Josef Fritz, Tanja Stocks

**Author notes:** Corresponding author: Anton Nilsson. Address: Register-based Epidemiology, CRC, Jan Waldenströms gata 35, SE-21428 Malmö, Sweden.

## Abstract

Excess body weight has been associated with increased cancer risk, but the role of weight change across adulthood remains unclear. We examined body weight trajectories from ages 17 to 60 and their associations with site-specific cancer incidence. Data were based on the ODDS study, a pooled, nationwide cohort study in Sweden, with data on weight spanning 1911 to 2020, and cancer follow-up through 2023. Weight trajectories were estimated with linear mixed effects models in individuals with at least three weight measurements. Cox regressions estimated hazard ratios for associations between weight trajectories and established and potentially obesity-related cancers. Fifth versus first quintile of weight change was associated with many cancers, most strongly with esophageal adenocarcinoma in men (HR 2.25; 95% CI 1.66-3.04), liver cancer in men (HR 2.67; 95% CI 2.15-3.33), endometrial cancer in women (HR 3.78; 95% CI 3.09-4.61), and pituitary tumors in both sexes (men: HR 3.13 [95% CI 2.13-4.61]; women: HR 2.13 [95% CI 1.41-3.22]). Associations varied by sex and age. Heavier weight at age 17 years and earlier obesity onset were also associated with higher cancer incidence. These findings highlight the importance of a life-course approach to weight management and support sex- and age-targeted cancer prevention strategies.

## Introduction

Obesity affects one in eight individuals globally^1^ and is recognized as a leading cause of cancer.^2^ As highlighted by the International Agency for Research on Cancer (IARC),^3^ substantial evidence has linked excess body weight to cancers of the esophagus (adenocarcinoma), gastric cardia, colorectum, liver, gallbladder, pancreas, postmenopausal breast, endometrium, ovaries, kidneys, meninges, thyroid gland, and multiple myeloma.^2,4–6^ Evidence of links to other cancer forms exists as well.^7–9^ A causal role of high body weight in cancer development is supported by several plausible biological mechanisms^10^ and, for at least eight cancer forms, by evidence from Mendelian randomization studies.^11^

Most studies on body weight and cancer have focused on weight at a single point in time – typically in mid to late adulthood – or on weight changes between two points.^12–20^ Associations between life-course weight trajectories based on multiple weight assessments and the risk of developing cancer remain less explored.^21–23^ Since the timing and extent of weight changes vary across individuals, it may however be important to consider weight changes continuously rather than at one or two time points, and to consider the role of weight changes in different age intervals.

The objective of this study was to examine body weight trajectories from ages 17 to 60 and their association with different cancers. We considered cancers that are established obesity-related according to the IARC^3^ as well as additional ones that have been identified as potentially obesity-related in prior analyses of our cohort.^7^ We distinguished between weight at age 17 and subsequent changes, and between weight changes across age intervals (17-30, 31-44, and 45-60), to assess if earlier and later weight changes were differentially associated with cancer risk. In addition, we examined age of obesity (body mass index; BMI ≥30 kg/m^2^) onset as an alternative way of quantifying weight gain and its timing.

## Methods

### Data

We used data from the Obesity and Disease Development Sweden (ODDS) study.^67^ ODDS is a collection of Swedish cohorts with information on weight, height, and smoking assessed on or more occasions from age 17 or later. The data have been linked, via unique personal identifiers, to national registers containing information on cancer, mortality, and sociodemographics (year of birth, sex, national background, marital status, education, and migration), enabling longitudinal follow-up across cohorts.

Two ODDS cohorts were established by government agencies for administrative purposes (the National Medical Birth Register and the Military Conscription Register), one through occupational health examinations of construction workers (Bygghälsan), and others through population-based health examinations or research initiatives. The pooled cohort includes 2,165,048 males (3,530,952 weight observations) and 2,130,811 females (4,202,949 weight observations). Depending on the cohort, data on weight and height were either objectively measured (85%), self-reported at the time of measurement (8%), or retrospectively self-reported (7%). Data were recorded between 1963 and 2020, with retrospective measurements dating back to 1911.

In the National Medical Birth Register, weights are normally measured between week eight and ten of the gestation. However, due to changes in the measurement procedure, weights measured before 1992 were not fully comparable to weights measured later, giving rise to a discontinuity in average weights over time. We therefore followed previous literature and corrected earlier weight measurements based on the time trends in weight.^68^

Following the Swedish National Board of Health and Welfare^69^ and previous work on ODDS,^7^ cancers were classified using the International Classification of Diseases (ICD) codes, WHO/HS/CANC/24.1 (Swedish PAD codes) and ICD-O/2 and ICD-O/3 (Swedish SNOMED codes). All primary malignant cancers were considered. Additionally, some tumors were counted as cancers regardless of malignancy due to their harmful nature, including all brain and central nervous system tumors and all endocrine tumors except those of the thyroid. Diagnosis codes are given in eTable 1.

Study outcomes included any cancer; established obesity-related cancers; potentially obesity-related cancers; cancers not classified as established or potentially obesity-related; and specific established or potentially obesity-related cancer types with at least 250 events per sex during follow-up.

Our exclusion criteria are illustrated in eFigure 1. We excluded observations for the following reasons: having a personal identifier that for administrative reasons had been changed or re-used, missing data on height, education or marital status, implausible values for weight or height (weight: <30 or >200 kg; height: <100 or >230 cm), weight at an age >60 years, prevalent cancer, or if the individual was recorded as dead or emigrated at the time of observation. Moreover, observations with less than a year from the observation date to the end of follow-up were removed to reduce reverse causation. Cohorts contributing <1% of remaining observations were excluded. Individuals with less than three eligible weight measurements after these exclusions were excluded.

The final study sample comprised 251,041 males (1,194,071 weight observations) and 378,981 females (1,340,772 weight observations). The distribution of individuals across the cohorts is shown in eTable 2. Most (70%) of the included weight observations in men came from the Construction Workers Cohort, whereas most (67%) of those in women came from the National Medical Birth Register. Following these, the SIMPLER (Swedish Infrastructure for Medical Population-based Life-course and Environmental Research) was the largest cohort in both men and women (14% of observations in men and 11% of observations in women).

Baseline was defined as one year after the third included weight measurement. End of follow-up was defined as the date of the first cancer diagnosis, death, emigration or the end of 2023, whichever came first. A few cancers could only be identified from 1993 as ICD-O-2 was not available previously (eTable 1) and for these outcomes, the follow-up was set to begin in 1993 at the earliest. For breast cancer, women were followed from age 55 at the earliest. Since the proportion of individuals with missing data on smoking was substantial (10%), we did not exclude observations with missing data on this variable but instead created a three-category variable for smoking, with one category for missing (current smoker/not current smoker/missing).

### Statistical analysis

We used a two-stage approach to estimate associations between weight trajectories and cancer incidence,^70,71^ separately in men and women. In the first stage, we estimated linear mixed effects (LME) models of weight through ages 17-60, with age centered at 17. To capture the population trend in weight across adulthood, we included natural cubic splines of age (four knots at prespecified quantiles, as per Harrell’s recommendations^72^). In addition, the models included random slopes and intercepts representing individual deviations from the population trend, which were estimated via Best Linear Unbiased Prediction (BLUP). We adjusted for the mode of weight measurement (objective current, self-reported current, or self-reported historical), and, for women, whether the weight measurement was from the National Medical Birth Register, as early pregnancy could influence weight. The estimated random slopes were categorized into sex-specific quintiles of weight change.

In the second stage, we estimated Cox regression models for cancer incidence with weight change quintile as the exposure. The Cox models employed age as the timescale and stratification with respect to birth cohort^73^ (<1940, 1940-1949, 1950-1959, 1960-1969, 1970-1979, and ≥1980). All models were covariate adjusted for height to account for differences in body size; height was treated as a fixed covariate and defined as the modal recorded value across measurements. In addition, models were adjusted for weight at age 17 (i.e., the estimated random intercept), country of birth of the individual and their parents, smoking status and marital status at the start of follow-up, and highest attained education.

We also assessed whether weight at age 17, divided into quintiles, was associated with the outcomes (with and without adjustment for the weight change quintiles). In separate models, we examined how weight changes in different age intervals were associated with the incidence of established obesity-related cancers. For this purpose, we divided ages into early (ages 17-29), middle (30-44), and later (45-60) adulthood. Since population trajectories were roughly linear within these age intervals, we utilized LME models with linear splines, with knots at the boundaries defining these intervals. In the Cox regressions, we here replaced the ages 17-60 weight change quintiles with the estimated slopes of the age-group-specific splines. Models were adjusted for the estimated slopes in previous age intervals. Furthermore, we assessed the role of age of obesity onset (the first age with a predicted BMI ≥30 kg/m^2^), as divided into early, middle, later adulthood, or post-60/never.

Sensitivity analyses included: 1) omitting the adjustment for smoking, 2) adding individuals with only two weight assessments (and beginning follow-up one year after the *second* weight observation), 3) omitting individuals with negative predicted weight changes in the three age intervals, and 4) considering BMI change instead of weight change.

Analyses were conducted in Stata/MP version 19.0 (StataCorp LLC, College Station, Texas), with two-sided tests and significance thresholds of 0.05.

## Results

### Descriptive statistics

The median number of weight measurements was 5 (interquartile range; IQR 4-7) in men and 3 (IQR 3-4) in women, and the median annual weight change from age 17-60 was 0.4 kg/year in both sexes. Weight trajectories across ages 17-60, aggregated into quintiles of weight change, are shown in eFigure 2. Descriptive characteristics for quintiles 1 and 5 are presented in Table 1, and in eTables 3-4 for all quintiles as well as for individuals excluded due to missing or extreme data. Individuals with steeper weight gain tended to be born later, with somewhat younger ages at start of follow-up. The proportion of individuals with missing data on smoking was higher in women than men, particularly in the steeper weight change quintiles.

**Table 1.** Descriptive Characteristics of the First and Fifth Quintiles of the Weight Trajectories.

|  | Males |  | Females |  |
| --- | --- | --- | --- | --- |
| Weight trajectory quintile | 1 | 5 | 1 | 5 |
| N individuals | 50,209 | 50,208 | 75,797 | 75,796 |
| N weight observations | 250,817 | 242,352 | 285,117 | 270,273 |
| Age at baseline<sup>a</sup> (years), median (IQR) | 42<br>(34-53) | 38<br>(28-47) | 40<br>(34-51) | 34<br>(30-38) |
| Weight at the first observation (kg), median (IQR) | 71<br>(65-78) | 72<br>(65-80) | 58<br>(53-64) | 68<br>(60-78) |
| Weight at the third observation (kg), median (IQR) | 72<br>(66-78) | 86<br>(78-95) | 58<br>(54-63) | 83<br>(75-92) |
| Birth year |  |  |  |  |
| <1940 | 22,295<br>(44.4%) | 9,658<br>(19.2%) | 10,710<br>(14.1%) | 24,666<br>(6.5%) |
| 1940-1949 | 13,225<br>(26.3%) | 13,526<br>(26.9%) | 14,571<br>(19.2%) | 41,946<br>(11.1%) |
| 1950-1959 | 10,329<br>(20.6%) | 16,184<br>(32.2%) | 12,839<br>(16.9%) | 50,844<br>(13.4%) |
| 1960-1969 | 4,026<br>(8.0%) | 10,111<br>(20.1%) | 12,799<br>(16.9%) | 82,880<br>(21.9%) |
| 1970-1979 | 244<br>(0.5%) | 593<br>(1.2%) | 15,554<br>(20.5%) | 102,749<br>(27.1%) |
| ≥1980 | 90<br>(0.2%) | 136<br>(0.3%) | 9,324<br>(12.3%) | 75,896<br>(20.0%) |
| Height (cm), mean (SD) | 176 (7) | 180 (7) | 164 (6) | 166 (6) |
| Highest achieved education during follow-up |  |  |  |  |
| <9 years primary education | 17,932<br>(35.7%) | 10,095<br>(20.1%) | 7,177<br>(9.5%) | 26,164<br>(6.9%) |
| 9 years primary education | 3,845<br>(7.7%) | 5,483<br>(10.9%) | 5,722<br>(7.5%) | 30,194<br>(8.0%) |
| <3 years secondary education | 15,715<br>(31.3%) | 21,503<br>(42.8%) | 19,233<br>(25.4%) | 91,281<br>(24.1%) |
| ≥3 years secondary education | 6,359<br>(12.7%) | 6,724<br>(13.4%) | 11,259<br>(14.9%) | 72,592<br>(19.2%) |
| Tertiary education | 6,358<br>(12.7%) | 6,403<br>(12.8%) | 32,406<br>(42.8%) | 158,750<br>(41.9%) |
| Country of birth |  |  |  |  |
| Born in Sweden with both parents born in Sweden | 46,281<br>(92.2%) | 45,646<br>(90.9%) | 62,753<br>(82.8%) | 289,194<br>(76.3%) |
| Born in Sweden with one or two parents born abroad | 1,541<br>(3.1%) | 2,516<br>(5.0%) | 5,428<br>(7.2%) | 32,375<br>(8.5%) |
| Born abroad | 2,387<br>(4.8%) | 2,046<br>(4.1%) | 7,616<br>(10.0%) | 57,412<br>(15.1%) |
| Marital status<sup>b</sup> |  |  |  |  |
| Unmarried | 14,483<br>(28.8%) | 20,855<br>(41.5%) | 18,078<br>(23.9%) | 102,833<br>(27.1%) |
| Married | 31,980<br>(63.7%) | 25,640<br>(51.1%) | 47,820<br>(63.1%) | 237,659<br>(62.7%) |
| Divorced | 3,348<br>(6.7%) | 3,495<br>(7.0%) | 8,542<br>(11.3%) | 34,853<br>(9.2%) |
| Widowed/-er | 398<br>(0.8%) | 218<br>(0.4%) | 1,357<br>(1.8%) | 3,636<br>(1.0%) |
| Smoker<sup>b</sup> |  |  |  |  |
| Yes | 18,104<br>(36.1%) | 15,953<br>(31.8%) | 14,244<br>(18.8%) | 55,098<br>(14.5%) |
| No | 31,565<br>(62.9%) | 33,822<br>(67.4%) | 54,900<br>(72.4%) | 267,705<br>(70.6%) |
| Missing | 540<br>(1.1%) | 433<br>(0.9%) | 6,653<br>(8.8%) | 56,178<br>(14.8%) |
| Follow-up time from baseline<sup>a</sup> to end of follow-up (years), median (IQR) | 32<br>(22-39) | 33<br>(22-39) | 19<br>(11-29) | 16<br>(9-23) |
| Incident cancer during follow-up |  |  |  |  |
| Yes | 15,037<br>(29.9%) | 11,989<br>(23.9%) | 9,589<br>(12.7%) | 35,676<br>(9.4%) |
| No | 35,172<br>(70.1%) | 38,219<br>(76.1%) | 66,208<br>(87.3%) | 343,305<br>(90.6%) |
| Age at cancer diagnosis (years), median (IQR) | 71<br>(64-77) | 67<br>(61-74) | 66<br>(57-74) | 62<br>(51-71) |

IQR = interquartile range, SD = standard deviation.
<sup>a</sup> Baseline is defined as one year after the third weight observation.
<sup>b</sup> As determined at the third weight observation.

During a median follow-up of 33 years in men (IQR 23-39) and 18 years in women (IQR 10-25), 27% of men and 9% of women developed cancer (eTables 3-4). Established obesity-related cancers with at least 250 events per sex included esophageal adenocarcinoma, gastric cardia cancer, and liver cancer in men; postmenopausal breast, endometrial, ovarian, and thyroid cancer in women; and colon, rectal, and pancreatic cancer, renal cell carcinoma, meningioma, and multiple myeloma in both sexes.

### Weight change across ages 17-60

Steeper weight gain was associated with higher incidence of any cancer, as well as many established obesity-related cancers in both men (Figure 1) and women (Figure 2). Associations tended to exhibit dose-response patterns. In what follows, we comment on the results for the fifth weight change quintile compared with the first. The hazard ratio (HR) for any cancer was 1.07 (95% CI 1.04-1.09) in men and 1.17 (95% CI 1.13-1.22) in women, whereas for established obesity-related cancers, it was 1.46 (95% CI 1.38-1.54) in men and 1.43 (95% CI 1.36-1.51) in women.

**Figure 1.**
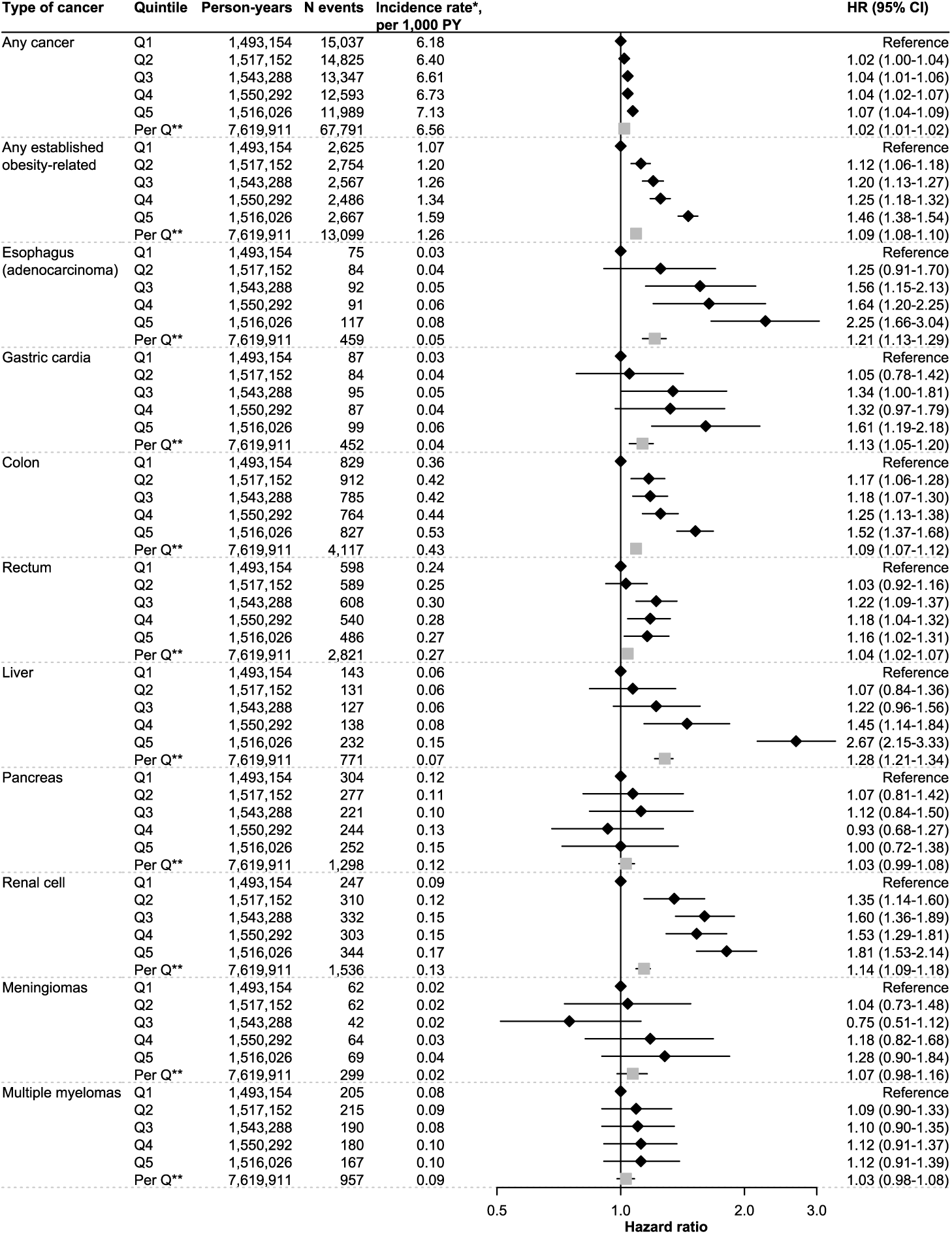
Any and Established Obesity-Related Cancers, by Weight Trajectory Quintile in Males. Hazard ratios (HRs) with 95% confidence intervals (CIs) for any, any established, and different sites of established obesity-related cancers by quintiles (Qs) of weight trajectories between ages 17 and 60. Models were estimated with Cox regression using age as the timescale, stratification with respect to birth cohort (<1940, 1940-1949, 1950-1959, 1960-1969, 1970-1979, and ≥1980), and covariate adjustment for predicted weight at age 17, height, country of birth of the individual and their parents, smoking, marital status, and the highest attained level of education. * standardized to the age distribution of the full Swedish population in 2020; ** effect of a one-quintile increase estimated from a linear model

**Figure 2.**
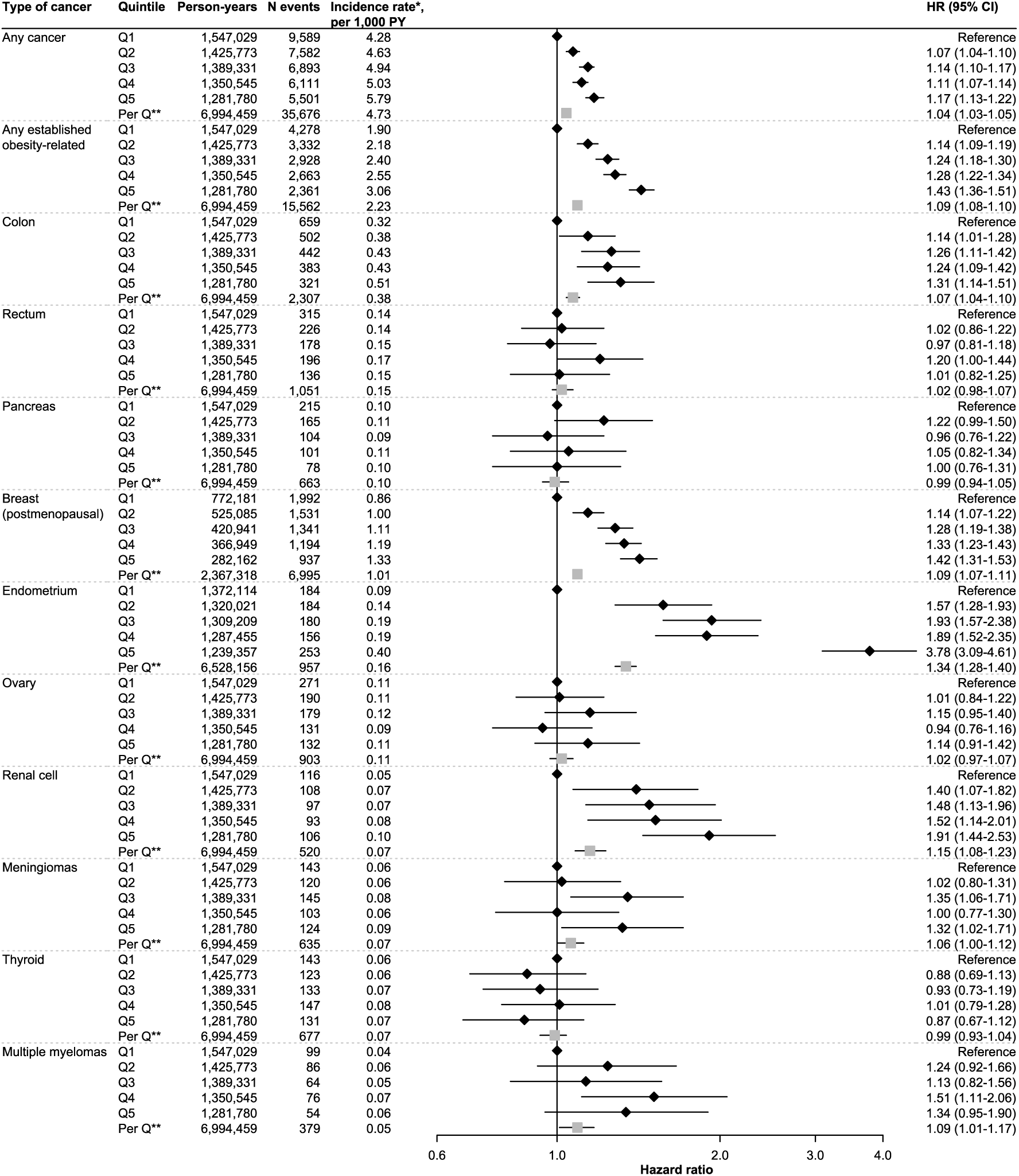
Any and Established Obesity-Related Cancers, by Weight Trajectory Quintile in Females. Hazard ratios (HRs) with 95% confidence intervals (CIs) for any, any established, and different sites of established obesity-related cancers by quintiles (Qs) of weight trajectories between ages 17 and 60. Models were estimated with Cox regression using age as the timescale, stratification with respect to birth cohort (<1940, 1940-1949, 1950-1959, 1960-1969, 1970-1979, and ≥1980), and covariate adjustment for predicted weight at age 17, height, country of birth of the individual and their parents, smoking, marital status, and the highest attained level of education. * standardized to the age distribution of the full Swedish population in 2020; ** effect of a one-quintile increase estimated from a linear model

For specific established obesity-related cancers, we detected substantial associations (HRs greater than 2) for liver cancer (HR 2.67; 95% CI 2.15-3.33) and esophageal adenocarcinoma (HR 2.25; 95% CI 1.66-3.04) in men, and for endometrial cancer (HR 3.78; 95% CI 3.09-4.61) in women. Statistically significant but weaker associations were seen for gastric cardia and rectal cancers in men, breast cancer and meningioma in women, and colon cancer and renal cell carcinoma in both men and women.

Among potentially obesity-related cancers (eFigures 3-4), substantial associations were seen for pituitary tumors in men (HR 3.13; 95% CI 2.13-4.61) and women (HR 2.13; 95% CI 1.41-3.22), with significant associations also for malignant melanoma and diffuse large B-cell lymphoma in men, and tumors of the parathyroid gland in women.

### Age 17 weight

Weight at age 17 was associated with several cancer types (eFigures 5-8). For most cancers, the HRs comparing the fifth and first quintiles of age 17 weight were comparable in magnitude to those for the fifth versus first quintile of weight change. Associations between age 17 weight and outcomes were virtually independent of subsequent weight change.

### Age-specific weight changes

Early, middle, and later adulthood weight changes were associated with overall cancer incidence and established obesity-related cancers in both men (Figure 3) and women (Figure 4). In women, associations per 0.5-kg weight increase were stronger for middle and later adulthood weight gain; in men, they were stronger for early adulthood weight gain.

**Figure 3.**
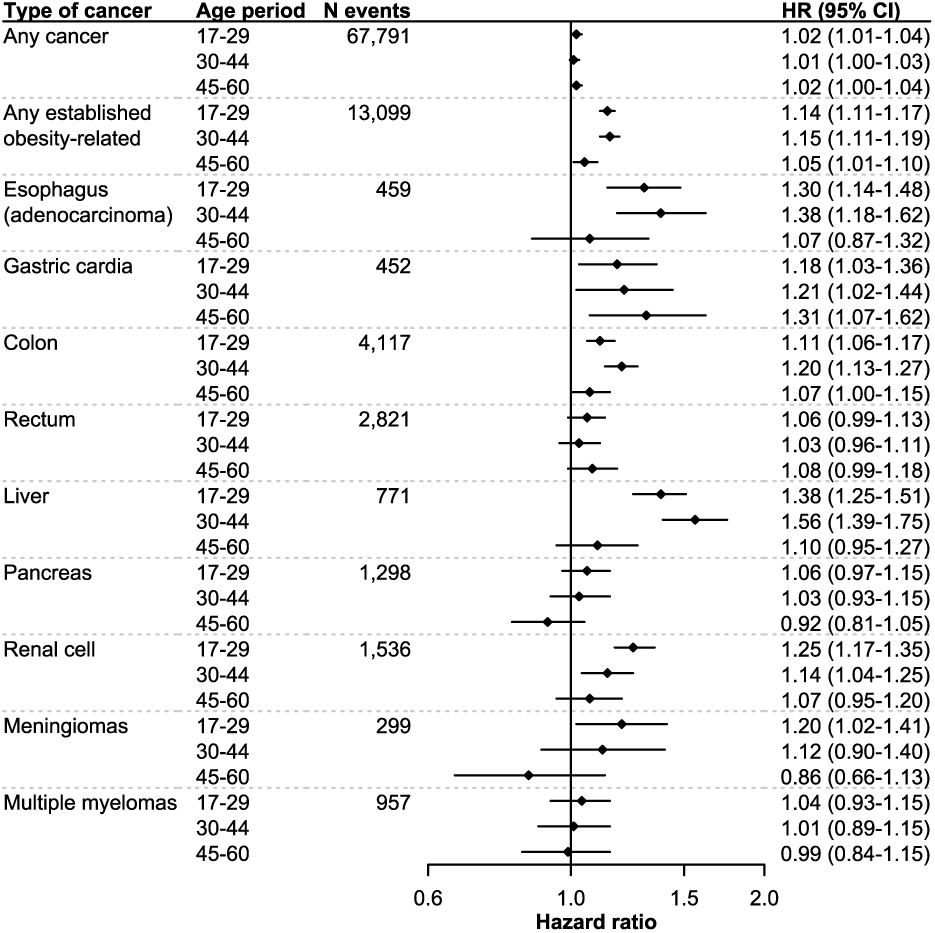
Any and Established Obesity-Related Cancers, by Age-Specific Weight Change in Males. Hazard ratios (HRs) with 95% confidence intervals (CIs) for any, established, and different sites of established obesity-related cancers per 0.5 kg weight increase per year. Models were estimated with Cox regression using age as the timescale, stratification with respect to birth cohort (<1940, 1940-1949, 1950-1959, 1960-1969, 1970-1979, and ≥1980), and covariate adjustment for predicted weight at age 17, height, country of birth of the individual and their parents, smoking, marital status, the highest attained level of education, and slopes of weight change in all previous age intervals.

**Figure 4.**
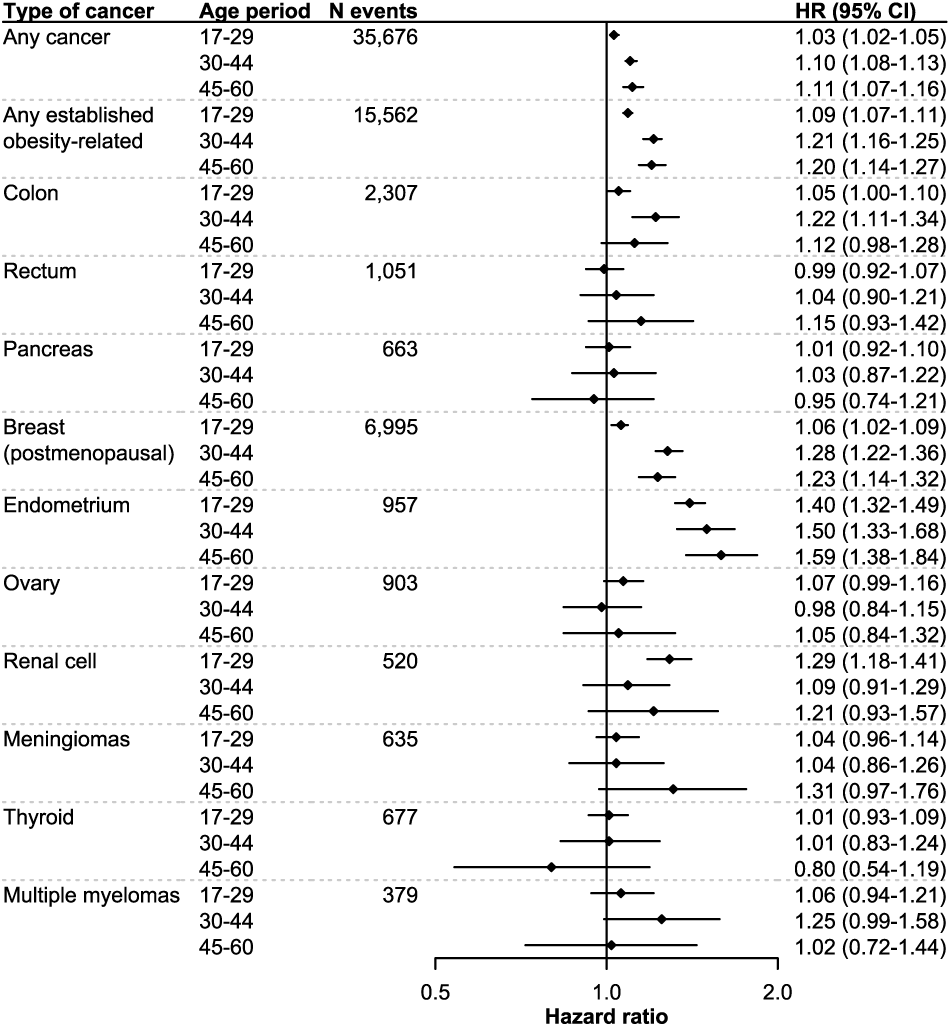
Any and Established Obesity-Related Cancers, by Age-Specific Weight Change in Females. Hazard ratios (HRs) with 95% confidence intervals (CIs) for any, established, and different sites of established obesity-related cancers per 0.5 kg weight increase per year. Models were estimated with Cox regression using age as the timescale, stratification with respect to birth cohort (<1940, 1940-1949, 1950-1959, 1960-1969, 1970-1979, and ≥1980), and covariate adjustment for predicted weight at age 17, height, country of birth of the individual and their parents, smoking, marital status, the highest attained level of education, and slopes of weight change in all previous age intervals.

To assess if this pattern in women was driven by female-specific cancers (breast, ovarian, endometrial), we conducted a secondary analysis excluding these cancers (eFigure 9). In this analysis, the associations for middle and later adulthood gain were more similar to those for early adulthood weight gain.

Some cancers were most strongly associated with early adulthood weight changes (particularly renal cell carcinoma in men), others with weight changes in middle adulthood (liver cancer in men and colon cancer in both sexes), and still others with later adulthood weight changes (gastric cardia cancer in men and meningioma in women). However, confidence intervals generally overlapped across age periods.

### Age of obesity onset

Analyses using age of obesity onset as the exposure (eFigures 10-11), revealed a tendency for cancer incidence to be more strongly associated with earlier ages of obesity onset. This pattern was particularly evident for liver and colon cancer in men, endometrial cancer in women, and renal cell carcinoma and pancreatic cancer in both men and women.

### Sensitivity analyses

In individuals with data on smoking, estimates were materially unchanged regardless of whether smoking was adjusted for (eFigures 12-13). Similarly robust results were obtained when we excluded individuals losing weight in an age interval (eFigures 14-15), added individuals with only two weight observations (eFigures 16-17), or compared quintiles of BMI change rather than weight change (eFigures 18-19).

## Discussion

Steeper increases in body weight between ages 17 and 60 were associated with higher incidence of several established obesity-related cancers, as well as some for which previous evidence linking obesity to cancer is more limited. Associations were particularly pronounced for liver cancer and esophageal adenocarcinoma in men, endometrial cancer in women, and renal cell carcinoma and pituitary tumors in both sexes.

Weight gain in women aged 30 years or older was strongly associated with endometrial cancer, postmenopausal breast cancer, and meningioma – cancers for which sex hormones are considered a primary etiological factor.^24–27^ Colon cancer was also strongly linked to female weight changes in these ages. In men, the associations with established obesity-related cancers were instead stronger for weight gains below age 45, most clearly for esophageal and liver cancer – cancers for which factors such as chronic inflammation, insulin resistance, and (in the case of esophageal cancer) gastroesophageal reflux disease (GERD) may play prominent roles.^28,29^ In general, the main biological mechanisms linking obesity to cancer are believed to include altered sex hormone metabolism, hyperinsulinemia/insulin-IGF signaling, and adipokine secretion and inflammation.^24,30–33^

Body weight at age 17 was independently associated with the risk of several cancers, with effects similar to or weaker than those observed for weight changes in ages 17-60. Notably, some cancers were associated with early adult weight but not with subsequent weight change, or vice versa. Pancreatic cancer, in particular, was in both sexes associated with weight at age 17 but not with weight change. Although pancreatic cancer is an established obesity-related cancer, prior studies have reported inconsistent associations with weight changes,^34–41^ with some evidence that early life body weight is particularly strongly related to the risk of developing this cancer.^42^

When weight increase was defined according to age at obesity onset, earlier onset tended to be more strongly associated with cancer incidence. Results for age of obesity onset are not directly comparable to those using quintiles of continuous measures of weight or weight change, but indicate that different operationalizations of adiposity may capture distinct dimensions of cancer risk.

Although numerous studies have reported links between body weight and cancer,^2–5,43–52^ only a few have examined how life-course weight trajectories relate to overall and site-specific cancer incidence.^21–23,53,54^ These studies have primarily focused on overweight duration and cumulative overweight. Our study extends this literature by focusing on weight change through adulthood and examining an unusually broad range of cancer sites. By distinguishing between initial adult body weight and subsequent weight change, and allowing weight dynamics to vary across age intervals, our analyses provide a more nuanced understanding of how adiposity over the life course relates to cancer risk.

Our results reinforce evidence suggesting that endometrial cancer is the cancer most strongly associated with weight gain^21,22,53,54^, a pattern also seen in studies using single BMI measurements.^2,3,5,7,10,55^ The strong associations between weight gain and liver cancer, esophageal adenocarcinoma, and renal cell carcinoma also confirm prior literature, although exact patterns have varied. To our knowledge, previous studies have not investigated weight changes as a risk factor for rare neoplasms such as pituitary tumors. A few studies used a single body weight measurement and documented moderate associations between obesity and pituitary tumors.^7,56,57^ Overall, while inherited genetic syndromes are major risk factors for pituitary tumors, evidence on the role of environmental factors is sparse.^58^

The strengths of this study include the large study population and long follow-up with repeated measurements, enabling precise estimation of associations across a wide range of cancer sites. Moreover, with most observations derived from the population-wide Medical Birth Register and the Construction Workers Cohort (participation rate 80%^59^), representativeness with respect to the background population is likely high. Although the Medical Birth Register is limited to women who have given birth, weight-cancer associations in ODDS have been found to be similar when excluding the Medical Birth Register.^7^ Moreover, the Swedish Cancer Register has high completeness and data quality, with 99% of all cancers morphologically verified.^60,61^ The observation that most established obesity-related cancers were positively associated with weight gain and higher initial weight provides an indication that results are not spurious. The only negative association – between higher weight at age 17 and lower postmenopausal breast cancer risk – is consistent with previous evidence.^41,62,63^

The study also has some limitations. We were unable to adjust for many confounding factors, including diet, physical activity, and genetic predisposition. However, adjustment for smoking had minimal impact on the results. We also lacked data on whether weight loss was intentional; however, removing individuals with weight loss made little difference. Moreover, given the etiological focus of our study, we did not apply corrections for multiple testing. It is therefore possible that some findings may be due to chance, and the results should be interpreted with some caution and replicated in future studies.

In summary, both early adult body weight and weight gain across adulthood were associated with the risk of most established and some potentially obesity-related cancers, with heterogeneity by cancer site, sex, and timing of weight gain. In the context of the rising prevalence of obesity^64,65^ and cancer^66^ in Western countries and globally, the findings highlight the importance of a life-course perspective on weight management for cancer prevention.

## Data Availability

The paper is based on administrative register data and other cohorts that are part of the ODDS study. Data access that is covered by the ethical approval will be considered in agreement with the principal investigator of ODDS, Tanja Stocks, and upon approval from the register holders and the steering committees of the ODDS cohorts.

## Author contributions

Dr Nilsson had full access to all data in the study and takes full responsibility for the integrity of the data and the accuracy of the data analysis.

## Concept and design

Stocks, da Silva, Le, Fritz, Nilsson

## Acquisition and preparation of data

Stocks, da Silva, Le, Häggström, Wahlström, Michaëlsson, Trolle Lagerros, Sandin, Magnusson, Fritz, Nilsson

**Drafting of the manuscript:** Nilsson

**Critical review of the manuscript for important intellectual content:** All

**Statistical analysis:** Nilsson

**Obtained funding:** Stocks

## Data sharing statement

All data are located on Statistics Sweden’s Microdata Online Access (MONA) server and may only be accessed from countries in the European Union or the European Economic Area. Data access covered by the ethical permit will be considered in agreement with the PI Tanja Stocks, and upon approval from register holders and steering committees of the ODDS cohorts.

## Supplementary Material for

**eTable 1.**
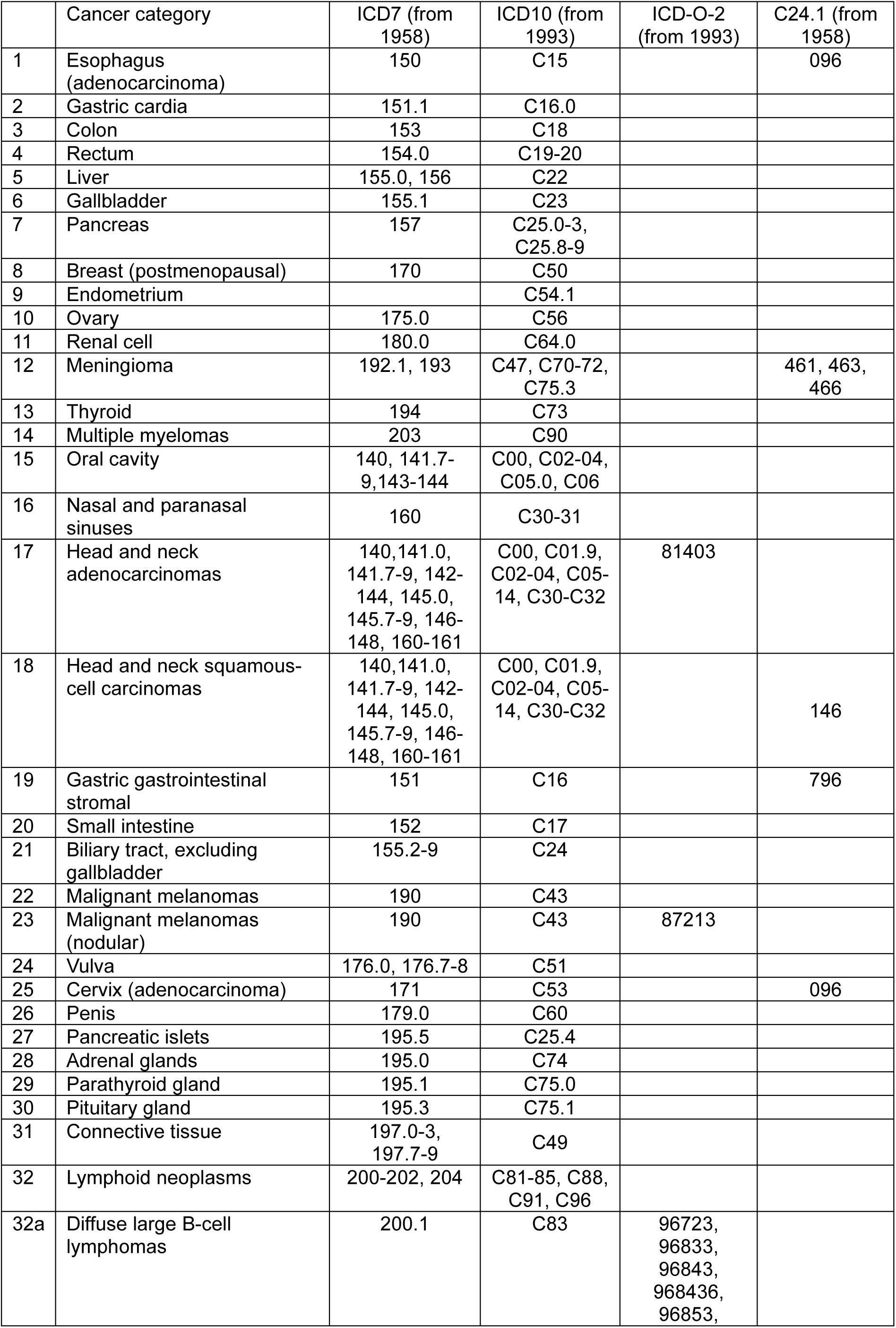
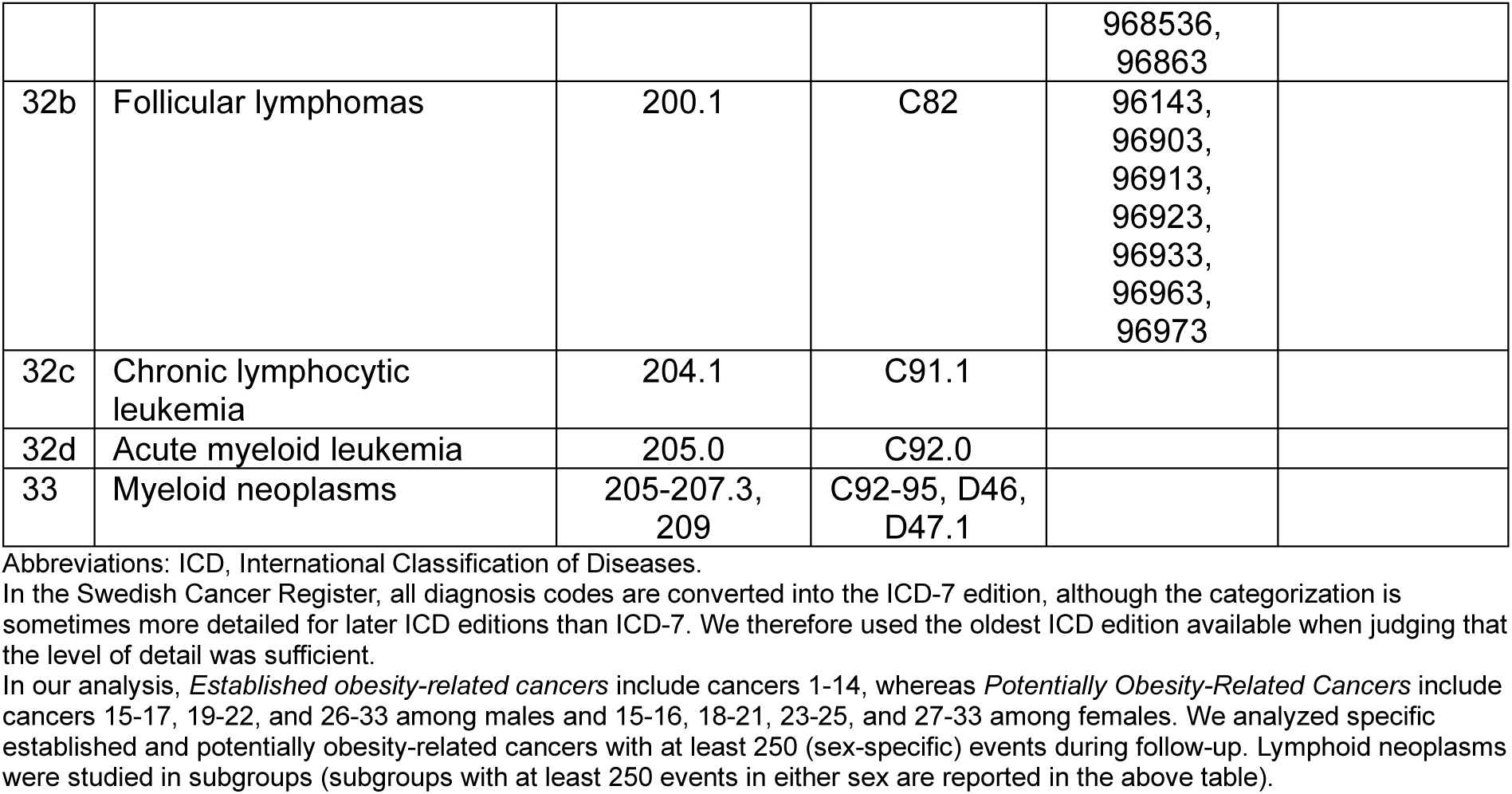
Cancer Diagnosis Codes.

|  | Cancer category | ICD7 (from 1958) | ICD10 (from 1993) | ICD-O-2 (from 1993) | C24.1 (from 1958) |
| --- | --- | --- | --- | --- | --- |
| 1 | Esophagus (adenocarcinoma) | 150 | C15 |  | 096 |
| 2 | Gastric cardia | 151.1 | C16.0 |  |  |
| 3 | Colon | 153 | C18 |  |  |
| 4 | Rectum | 154.0 | C19-20 |  |  |
| 5 | Liver | 155.0, 156 | C22 |  |  |
| 6 | Gallbladder | 155.1 | C23 |  |  |
| 7 | Pancreas | 157 | C25.0-3, C25.8-9 |  |  |
| 8 | Breast (postmenopausal) | 170 | C50 |  |  |
| 9 | Endometrium |  | C54.1 |  |  |
| 10 | Ovary | 175.0 | C56 |  |  |
| 11 | Renal cell | 180.0 | C64.0 |  |  |
| 12 | Meningioma | 192.1, 193 | C47, C70-72, C75.3 |  | 461, 463, 466 |
| 13 | Thyroid | 194 | C73 |  |  |
| 14 | Multiple myelomas | 203 | C90 |  |  |
| 15 | Oral cavity | 140, 141.7-9, 143-144 | C00, C02-04, C05.0, C06 |  |  |
| 16 | Nasal and paranasal sinuses | 160 | C30-31 |  |  |
| 17 | Head and neck adenocarcinomas | 140, 141.0, 141.7-9, 142-144, 145.0, 145.7-9, 146-148, 160-161 | C00, C01.9, C02-04, C05-14, C30-C32 | 81403 |  |
| 18 | Head and neck squamous-cell carcinomas | 140, 141.0, 141.7-9, 142-144, 145.0, 145.7-9, 146-148, 160-161 | C00, C01.9, C02-04, C05-14, C30-C32 |  | 146 |
| 19 | Gastric gastrointestinal stromal | 151 | C16 |  | 796 |
| 20 | Small intestine | 152 | C17 |  |  |
| 21 | Biliary tract, excluding gallbladder | 155.2-9 | C24 |  |  |
| 22 | Malignant melanomas | 190 | C43 |  |  |
| 23 | Malignant melanomas (nodular) | 190 | C43 | 87213 |  |
| 24 | Vulva | 176.0, 176.7-8 | C51 |  |  |
| 25 | Cervix (adenocarcinoma) | 171 | C53 |  | 096 |
| 26 | Penis | 179.0 | C60 |  |  |
| 27 | Pancreatic islets | 195.5 | C25.4 |  |  |
| 28 | Adrenal glands | 195.0 | C74 |  |  |
| 29 | Parathyroid gland | 195.1 | C75.0 |  |  |
| 30 | Pituitary gland | 195.3 | C75.1 |  |  |
| 31 | Connective tissue | 197.0-3, 197.7-9 | C49 |  |  |
| 32 | Lymphoid neoplasms | 200-202, 204 | C81-85, C88, C91, C96 |  |  |
| 32a | Diffuse large B-cell lymphomas | 200.1 | C83 | 96723, 96833, 96843, 968436, 96853, |  |
|  |  |  |  | 968536,<br>96863 |  |
| 32b | Follicular lymphomas | 200.1 | C82 | 96143,<br>96903,<br>96913,<br>96923,<br>96933,<br>96963,<br>96973 |  |
| 32c | Chronic lymphocytic leukemia | 204.1 | C91.1 |  |  |
| 32d | Acute myeloid leukemia | 205.0 | C92.0 |  |  |
| 33 | Myeloid neoplasms | 205-207.3,<br>209 | C92-95, D46,<br>D47.1 |  |  |

Abbreviations: ICD, International Classification of Diseases.
In the Swedish Cancer Register, all diagnosis codes are converted into the ICD-7 edition, although the categorization is sometimes more detailed for later ICD editions than ICD-7. We therefore used the oldest ICD edition available when judging that the level of detail was sufficient.
In our analysis, *Established obesity-related cancers* include cancers 1-14, whereas *Potentially Obesity-Related Cancers* include cancers 15-17, 19-22, and 26-33 among males and 15-16, 18-21, 23-25, and 27-33 among females. We analyzed specific established and potentially obesity-related cancers with at least 250 (sex-specific) events during follow-up. Lymphoid neoplasms were studied in subgroups (subgroups with at least 250 events in either sex are reported in the above table).

**eTable 2.** Number and Years of Weight Observations per Cohort in the Study Sample.

| Cohort | Number of observations among males | Number of observations among females | Years of data collection on body weight |
| --- | --- | --- | --- |
| National Medical Birth Register | - | 895,807 | 1982-2019 |
| Swedish Military Conscription Register | 91,498 | 6,232 | 1969-2014 |
| Construction Workers Cohort | 838,012 | 27,313 | 1971-1993 |
| Malmö cohorts* | 17,280 | 16,855 | 1974-2019 |
| Swedish National March Cohort | 14,969 | 29,732 | 1997 |
| Northern Sweden Health and Disease Study | 49,911 | 83,728 | 1985-2020 |
| SIMPLER | 163,369 | 154,106 | 1987-2008 |
| Swedish Twin Registry | 19,032 | 28,002 | 1963-2013 |
| Women's Lifestyle and Health | - | 98,997 | 1991-2004 |
| Total | 1,194,071 | 1,340,772 | 1963-2020 |

\*Includes the Malmö Preventive Project, the Malmö Diet and Cancer Study, and the Malmö Offspring Study

**eTable 3.**
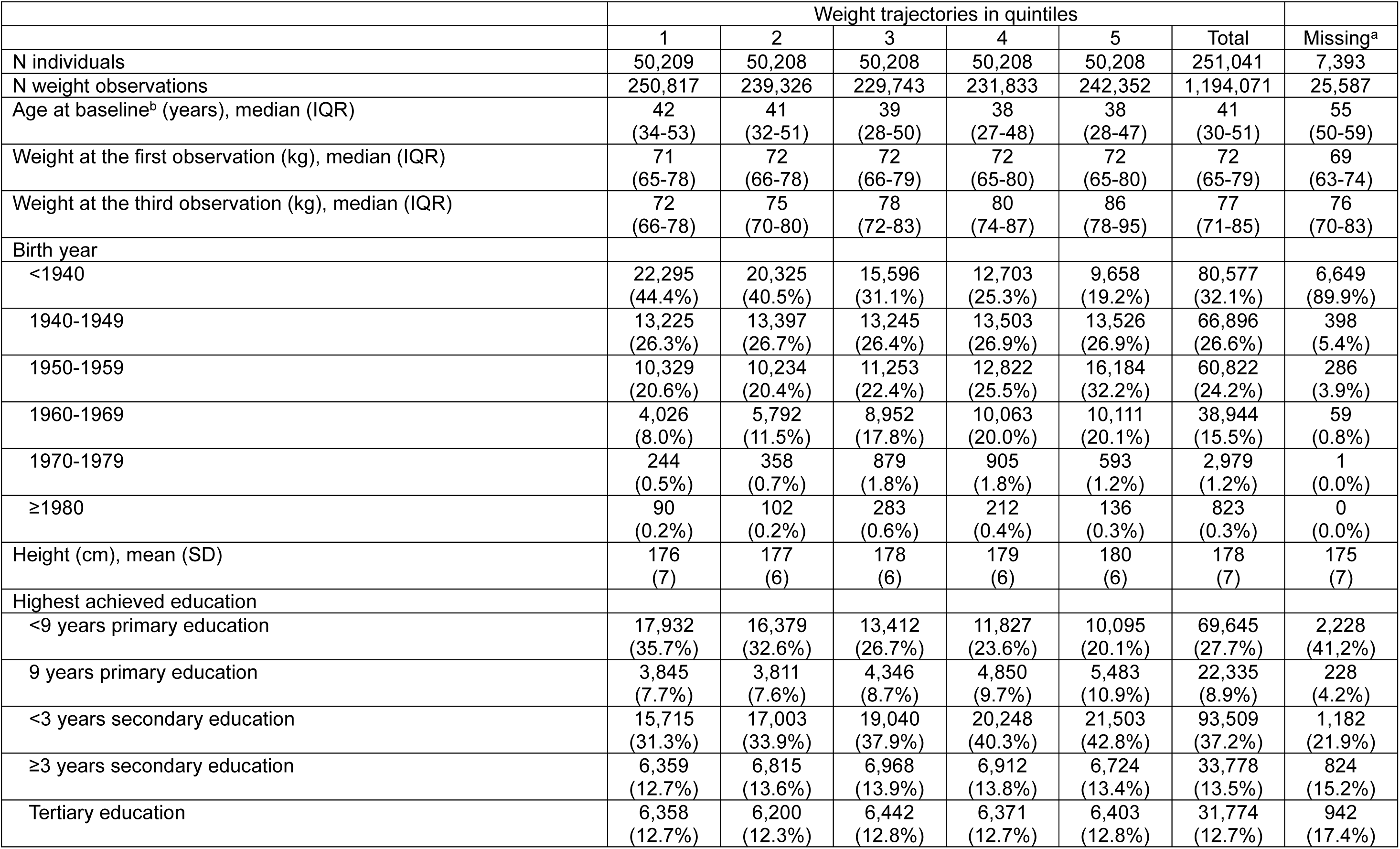
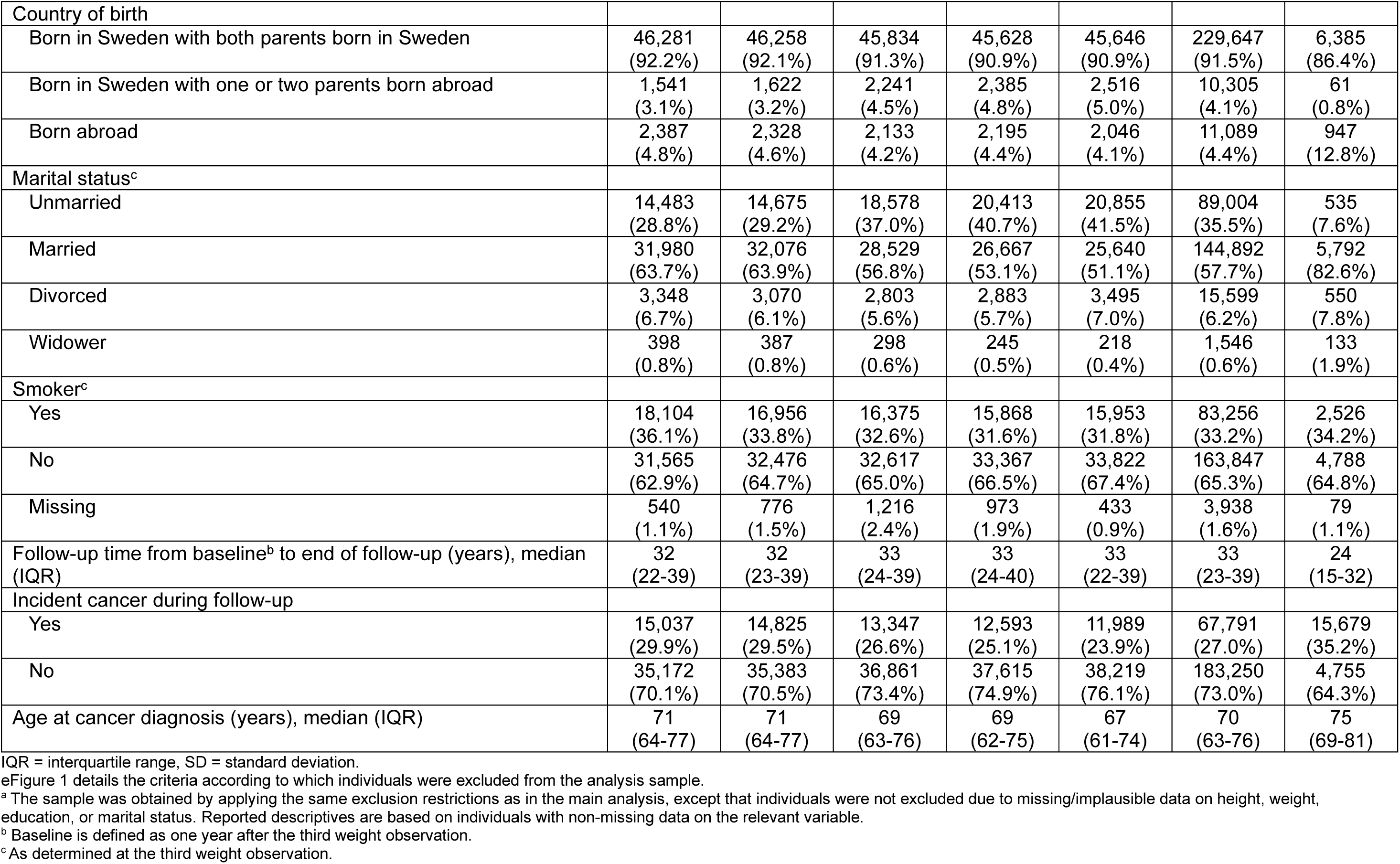
Descriptive Characteristics, Males.

|  | Weight trajectories in quintiles |  |  |  |  |  | Missing<sup>a</sup> |
| --- | --- | --- | --- | --- | --- | --- | --- |
|  | 1 | 2 | 3 | 4 | 5 | Total |  |
| N individuals | 50,209 | 50,208 | 50,208 | 50,208 | 50,208 | 251,041 | 7,393 |
| N weight observations | 250,817 | 239,326 | 229,743 | 231,833 | 242,352 | 1,194,071 | 25,587 |
| Age at baseline<sup>b</sup> (years), median (IQR) | 42<br>(34-53) | 41<br>(32-51) | 39<br>(28-50) | 38<br>(27-48) | 38<br>(28-47) | 41<br>(30-51) | 55<br>(50-59) |
| Weight at the first observation (kg), median (IQR) | 71<br>(65-78) | 72<br>(66-78) | 72<br>(66-79) | 72<br>(65-80) | 72<br>(65-80) | 72<br>(65-79) | 69<br>(63-74) |
| Weight at the third observation (kg), median (IQR) | 72<br>(66-78) | 75<br>(70-80) | 78<br>(72-83) | 80<br>(74-87) | 86<br>(78-95) | 77<br>(71-85) | 76<br>(70-83) |
| Birth year |  |  |  |  |  |  |  |
| <1940 | 22,295<br>(44.4%) | 20,325<br>(40.5%) | 15,596<br>(31.1%) | 12,703<br>(25.3%) | 9,658<br>(19.2%) | 80,577<br>(32.1%) | 6,649<br>(89.9%) |
| 1940-1949 | 13,225<br>(26.3%) | 13,397<br>(26.7%) | 13,245<br>(26.4%) | 13,503<br>(26.9%) | 13,526<br>(26.9%) | 66,896<br>(26.6%) | 398<br>(5.4%) |
| 1950-1959 | 10,329<br>(20.6%) | 10,234<br>(20.4%) | 11,253<br>(22.4%) | 12,822<br>(25.5%) | 16,184<br>(32.2%) | 60,822<br>(24.2%) | 286<br>(3.9%) |
| 1960-1969 | 4,026<br>(8.0%) | 5,792<br>(11.5%) | 8,952<br>(17.8%) | 10,063<br>(20.0%) | 10,111<br>(20.1%) | 38,944<br>(15.5%) | 59<br>(0.8%) |
| 1970-1979 | 244<br>(0.5%) | 358<br>(0.7%) | 879<br>(1.8%) | 905<br>(1.8%) | 593<br>(1.2%) | 2,979<br>(1.2%) | 1<br>(0.0%) |
| ≥1980 | 90<br>(0.2%) | 102<br>(0.2%) | 283<br>(0.6%) | 212<br>(0.4%) | 136<br>(0.3%) | 823<br>(0.3%) | 0<br>(0.0%) |
| Height (cm), mean (SD) | 176<br>(7) | 177<br>(6) | 178<br>(6) | 179<br>(6) | 180<br>(6) | 178<br>(7) | 175<br>(7) |
| Highest achieved education |  |  |  |  |  |  |  |
| <9 years primary education | 17,932<br>(35.7%) | 16,379<br>(32.6%) | 13,412<br>(26.7%) | 11,827<br>(23.6%) | 10,095<br>(20.1%) | 69,645<br>(27.7%) | 2,228<br>(41.2%) |
| 9 years primary education | 3,845<br>(7.7%) | 3,811<br>(7.6%) | 4,346<br>(8.7%) | 4,850<br>(9.7%) | 5,483<br>(10.9%) | 22,335<br>(8.9%) | 228<br>(4.2%) |
| <3 years secondary education | 15,715<br>(31.3%) | 17,003<br>(33.9%) | 19,040<br>(37.9%) | 20,248<br>(40.3%) | 21,503<br>(42.8%) | 93,509<br>(37.2%) | 1,182<br>(21.9%) |
| ≥3 years secondary education | 6,359<br>(12.7%) | 6,815<br>(13.6%) | 6,968<br>(13.9%) | 6,912<br>(13.8%) | 6,724<br>(13.4%) | 33,778<br>(13.5%) | 824<br>(15.2%) |
| Tertiary education | 6,358<br>(12.7%) | 6,200<br>(12.3%) | 6,442<br>(12.8%) | 6,371<br>(12.7%) | 6,403<br>(12.8%) | 31,774<br>(12.7%) | 942<br>(17.4%) |
| Country of birth |  |  |  |  |  |  |  |
| Born in Sweden with both parents born in Sweden | 46,281<br>(92.2%) | 46,258<br>(92.1%) | 45,834<br>(91.3%) | 45,628<br>(90.9%) | 45,646<br>(90.9%) | 229,647<br>(91.5%) | 6,385<br>(86.4%) |
| Born in Sweden with one or two parents born abroad | 1,541<br>(3.1%) | 1,622<br>(3.2%) | 2,241<br>(4.5%) | 2,385<br>(4.8%) | 2,516<br>(5.0%) | 10,305<br>(4.1%) | 61<br>(0.8%) |
| Born abroad | 2,387<br>(4.8%) | 2,328<br>(4.6%) | 2,133<br>(4.2%) | 2,195<br>(4.4%) | 2,046<br>(4.1%) | 11,089<br>(4.4%) | 947<br>(12.8%) |
| Marital status<sup>c</sup> |  |  |  |  |  |  |  |
| Unmarried | 14,483<br>(28.8%) | 14,675<br>(29.2%) | 18,578<br>(37.0%) | 20,413<br>(40.7%) | 20,855<br>(41.5%) | 89,004<br>(35.5%) | 535<br>(7.6%) |
| Married | 31,980<br>(63.7%) | 32,076<br>(63.9%) | 28,529<br>(56.8%) | 26,667<br>(53.1%) | 25,640<br>(51.1%) | 144,892<br>(57.7%) | 5,792<br>(82.6%) |
| Divorced | 3,348<br>(6.7%) | 3,070<br>(6.1%) | 2,803<br>(5.6%) | 2,883<br>(5.7%) | 3,495<br>(7.0%) | 15,599<br>(6.2%) | 550<br>(7.8%) |
| Widower | 398<br>(0.8%) | 387<br>(0.8%) | 298<br>(0.6%) | 245<br>(0.5%) | 218<br>(0.4%) | 1,546<br>(0.6%) | 133<br>(1.9%) |
| Smoker<sup>c</sup> |  |  |  |  |  |  |  |
| Yes | 18,104<br>(36.1%) | 16,956<br>(33.8%) | 16,375<br>(32.6%) | 15,868<br>(31.6%) | 15,953<br>(31.8%) | 83,256<br>(33.2%) | 2,526<br>(34.2%) |
| No | 31,565<br>(62.9%) | 32,476<br>(64.7%) | 32,617<br>(65.0%) | 33,367<br>(66.5%) | 33,822<br>(67.4%) | 163,847<br>(65.3%) | 4,788<br>(64.8%) |
| Missing | 540<br>(1.1%) | 776<br>(1.5%) | 1,216<br>(2.4%) | 973<br>(1.9%) | 433<br>(0.9%) | 3,938<br>(1.6%) | 79<br>(1.1%) |
| Follow-up time from baseline<sup>b</sup> to end of follow-up (years), median (IQR) | 32<br>(22-39) | 32<br>(23-39) | 33<br>(24-39) | 33<br>(24-40) | 33<br>(22-39) | 33<br>(23-39) | 24<br>(15-32) |
| Incident cancer during follow-up |  |  |  |  |  |  |  |
| Yes | 15,037<br>(29.9%) | 14,825<br>(29.5%) | 13,347<br>(26.6%) | 12,593<br>(25.1%) | 11,989<br>(23.9%) | 67,791<br>(27.0%) | 15,679<br>(35.2%) |
| No | 35,172<br>(70.1%) | 35,383<br>(70.5%) | 36,861<br>(73.4%) | 37,615<br>(74.9%) | 38,219<br>(76.1%) | 183,250<br>(73.0%) | 4,755<br>(64.3%) |
| Age at cancer diagnosis (years), median (IQR) | 71<br>(64-77) | 71<br>(64-77) | 69<br>(63-76) | 69<br>(62-75) | 67<br>(61-74) | 70<br>(63-76) | 75<br>(69-81) |

IQR = interquartile range, SD = standard deviation.
<sup>e</sup>Figure 1 details the criteria according to which individuals were excluded from the analysis sample.
<sup>a</sup> The sample was obtained by applying the same exclusion restrictions as in the main analysis, except that individuals were not excluded due to missing/improbable data on height, weight, education, or marital status. Reported descriptives are based on individuals with non-missing data on the relevant variable.
<sup>b</sup> Baseline is defined as one year after the third weight observation.
<sup>c</sup> As determined at the third weight observation.

**eTable 4.** Descriptive Characteristics, Females.

|  | Weight trajectories in quintiles |  |  |  |  |  | Missing<sup>a</sup> |
| --- | --- | --- | --- | --- | --- | --- | --- |
|  | 1 | 2 | 3 | 4 | 5 | Total |  |
| N individuals | 75,797 | 75,796 | 75,796 | 75,796 | 75,796 | 378,981 | 9,672 |
| N weight observations | 285,117 | 264,656 | 259,553 | 261,173 | 270,273 | 1,340,772 | 32,771 |
| Age at baseline<sup>b</sup> (years), median (IQR) | 40<br>(34-51) | 36<br>(32-41) | 35<br>(31-40) | 34<br>(31-39) | 34<br>(30-38) | 35<br>(31-41) | 53<br>(41-60) |
| Weight at the first observation (kg), median (IQR) | 58<br>(53-64) | 59<br>(54-64) | 61<br>(56-66) | 63<br>(57-70) | 68<br>(60-78) | 61<br>(55-68) | 56<br>(51-62) |
| Weight at the third observation (kg), median (IQR) | 58<br>(54-63) | 61<br>(57-65) | 65<br>(61-70) | 71<br>(67-76) | 83<br>(75-92) | 66<br>(60-75) | 65<br>(59-73) |
| Birth year |  |  |  |  |  |  |  |
| <1940 | 10,710<br>(14.1%) | 5,836<br>(7.7%) | 3,780<br>(5.0%) | 2,729<br>(3.6%) | 1,611<br>(2.1%) | 24,666<br>(6.5%) | 6,250<br>(64.6%) |
| 1940-1949 | 14,571<br>(19.2%) | 9,290<br>(12.3%) | 7,181<br>(9.5%) | 6,297<br>(8.3%) | 4,607<br>(6.1%) | 41,946<br>(11.1%) | 840<br>(8.7%) |
| 1950-1959 | 12,839<br>(16.9%) | 11,036<br>(14.6%) | 10,141<br>(13.4%) | 9,233<br>(12.2%) | 7,595<br>(10.0%) | 50,844<br>(13.4%) | 574<br>(5.9%) |
| 1960-1969 | 12,799<br>(16.9%) | 15,995<br>(21.1%) | 17,428<br>(23.0%) | 17,945<br>(23.7%) | 18,713<br>(24.7%) | 82,880<br>(21.9%) | 514<br>(5.3%) |
| 1970-1979 | 15,554<br>(20.5%) | 20,206<br>(26.7%) | 21,314<br>(28.1%) | 21,772<br>(28.7%) | 23,903<br>(31.5%) | 102,749<br>(27.1%) | 611<br>(6.3%) |
| ≥1980 | 9,324<br>(12.3%) | 13,433<br>(17.7%) | 15,952<br>(21.0%) | 17,820<br>(23.5%) | 19,367<br>(25.6%) | 75,896<br>(20.0%) | 883<br>(9.1%) |
| Height (cm), mean (SD) | 164<br>(6) | 165<br>(6) | 166<br>(6) | 167<br>(6) | 167<br>(6) | 166<br>(6) | 163<br>(6) |
| Highest achieved education |  |  |  |  |  |  |  |
| <9 years primary education | 7,177<br>(9.5%) | 4,807<br>(6.3%) | 4,162<br>(5.5%) | 4,656<br>(6.1%) | 5,362<br>(7.1%) | 26,164<br>(6.9%) | 2,574<br>(37.4%) |
| 9 years primary education | 5,722<br>(7.5%) | 5,036<br>(6.6%) | 5,240<br>(6.9%) | 6,097<br>(8.0%) | 8,099<br>(10.7%) | 30,194<br>(8.0%) | 591<br>(8.6%) |
| <3 years secondary education | 19,233<br>(25.4%) | 16,681<br>(22.0%) | 16,893<br>(22.3%) | 18,157<br>(24.0%) | 20,317<br>(26.8%) | 91,281<br>(24.1%) | 1,799<br>(26.1%) |
| ≥3 years secondary education | 11,259<br>(14.9%) | 12,941<br>(17.1%) | 14,854<br>(19.6%) | 15,898<br>(21.0%) | 17,640<br>(23.3%) | 72,592<br>(19.2%) | 523<br>(7.6%) |
| Tertiary education | 32,406<br>(42.8%) | 36,331<br>(47.9%) | 34,647<br>(45.7%) | 30,988<br>(40.9%) | 24,378<br>(32.2%) | 158,750<br>(41.9%) | 1,398<br>(20.3%) |
| Country of birth |  |  |  |  |  |  |  |
| Born in Sweden with both parents born in Sweden | 62,753<br>(82.8%) | 60,463<br>(79.8%) | 58,563<br>(77.3%) | 55,385<br>(73.1%) | 52,030<br>(68.6%) | 289,194<br>(76.3%) | 6,003<br>(62.1%) |
| Born in Sweden with one or two parents born abroad | 5,428<br>(7.2%) | 6,239<br>(8.2%) | 6,662<br>(8.8%) | 6,842<br>(9.0%) | 7,204<br>(9.5%) | 32,375<br>(8.5%) | 81<br>(0.8%) |
| Born abroad | 7,616<br>(10.0%) | 9,094<br>(12.0%) | 10,571<br>(13.9%) | 13,569<br>(17.9%) | 16,562<br>(21.9%) | 57,412<br>(15.1%) | 3,588<br>(37.1%) |
| Marital status<sup>c</sup> |  |  |  |  |  |  |  |
| Unmarried | 18,078<br>(23.9%) | 19,321<br>(25.5%) | 20,912<br>(27.6%) | 21,390<br>(28.2%) | 23,132<br>(30.5%) | 102,833<br>(27.1%) | 494<br>(5.4%) |
| Married | 47,820<br>(63.1%) | 49,286<br>(65.0%) | 48,426<br>(63.9%) | 47,570<br>(62.8%) | 44,557<br>(58.8%) | 237,659<br>(62.7%) | 6,916<br>(75.7%) |
| Divorced | 8,542<br>(11.3%) | 6,429<br>(8.5%) | 5,845<br>(7.7%) | 6,364<br>(8.4%) | 7,673<br>(10.1%) | 34,853<br>(9.2%) | 1,191<br>(13.0%) |
| Widowed | 1,357<br>(1.8%) | 760<br>(1.0%) | 613<br>(0.8%) | 472<br>(0.6%) | 434<br>(0.6%) | 3,636<br>(1.0%) | 541<br>(5.9%) |
| Smoker<sup>c</sup> |  |  |  |  |  |  |  |
| Yes | 14,244<br>(18.8%) | 9,907<br>(13.1%) | 9,495<br>(12.5%) | 10,106<br>(13.3%) | 11,346<br>(15.0%) | 55,098<br>(14.5%) | 1,535<br>(15.9%) |
| No | 54,900<br>(72.4%) | 54,771<br>(72.3%) | 53,092<br>(70.0%) | 52,663<br>(69.5%) | 52,279<br>(69.0%) | 267,705<br>(70.6%) | 7,624<br>(78.8%) |
| Missing | 6,653<br>(8.8%) | 11,118<br>(14.7%) | 13,209<br>(17.4%) | 13,027<br>(17.2%) | 12,171<br>(16.1%) | 56,178<br>(14.8%) | 513<br>(5.3%) |
| Follow-up time from baseline<sup>b</sup> to end of follow-up (years), median (IQR) | 19<br>(11-29) | 18<br>(10-26) | 17<br>(10-25) | 17<br>(10-25) | 16<br>(9-23) | 18<br>(10-25) | 23<br>(13-30) |
| Follow-up ending with cancer event |  |  |  |  |  |  |  |
| Yes | 9,589<br>(12.7%) | 7,582<br>(10.0%) | 6,892<br>(9.1%) | 6,112<br>(8.1%) | 5,501<br>(7.3%) | 35,676<br>(9.4%) | 2,136<br>(22.1%) |
| No | 66,208<br>(87.3%) | 68,214<br>(90.0%) | 68,904<br>(90.9%) | 69,684<br>(91.9%) | 70,295<br>(92.7%) | 343,305<br>(90.6%) | 7,536<br>(77.9%) |
| Age at cancer diagnosis, median (IQR) | 66<br>(57-74) | 62<br>(52-72) | 60<br>(49-69) | 59<br>(49-68) | 57<br>(48-66) | 62<br>(51-71) | 73<br>(65-81) |

IQR = interquartile range, SD = standard deviation.
<sup>e</sup>Figure 1 details the criteria according to which individuals were excluded from the analysis sample.
<sup>a</sup> The sample was obtained by applying the same exclusion restrictions as in the main analysis, except that individuals were not excluded due to missing/improbable data on height, weight, education, or marital status. Reported descriptives are based on individuals with non-missing data on the relevant variable.
<sup>b</sup> Baseline is defined as one year after the third weight observation.
<sup>c</sup> As determined at the third weight observation.

**eFigure 1.**
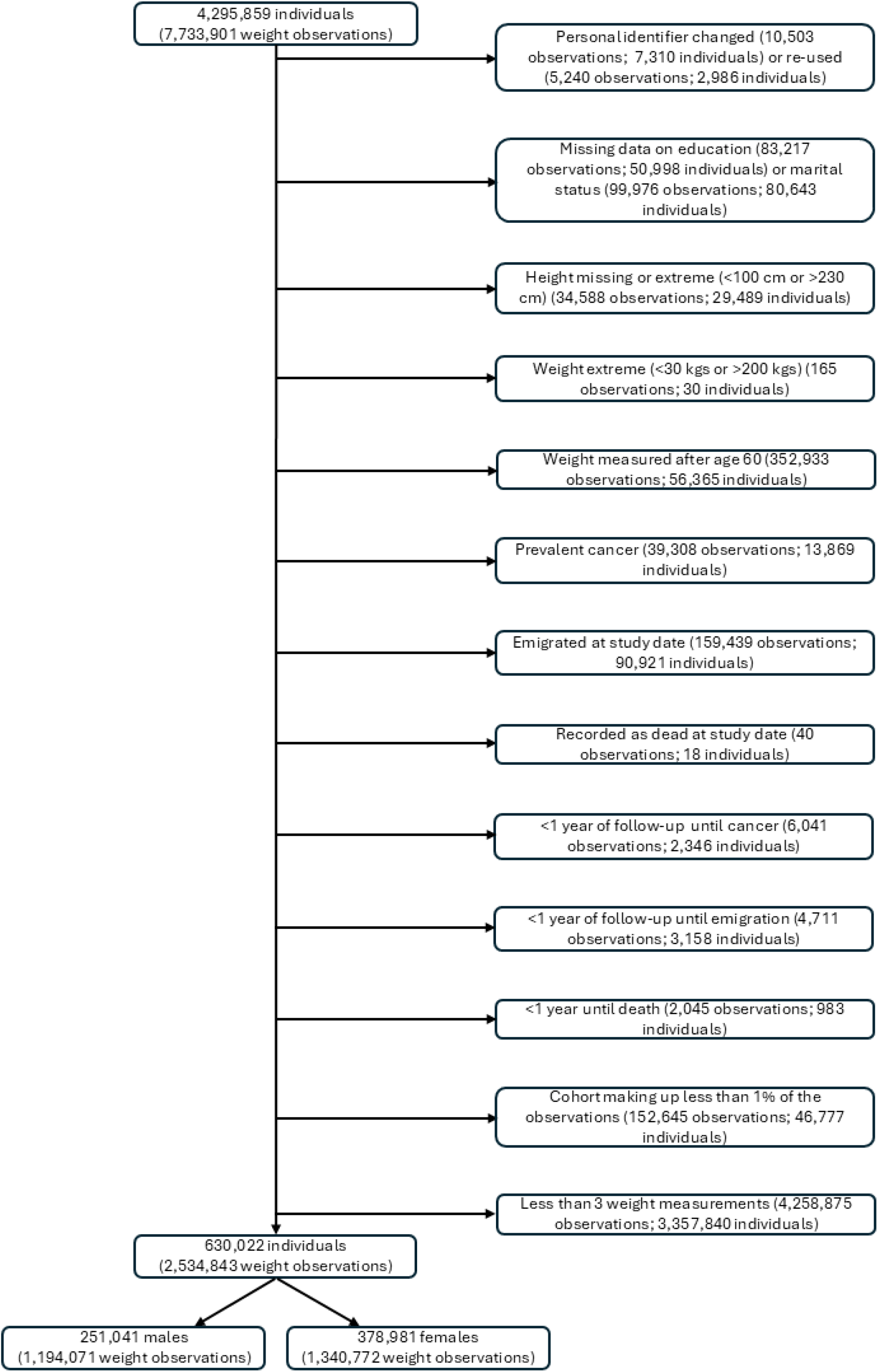
Exclusion Criteria.

**eFigure 2.**
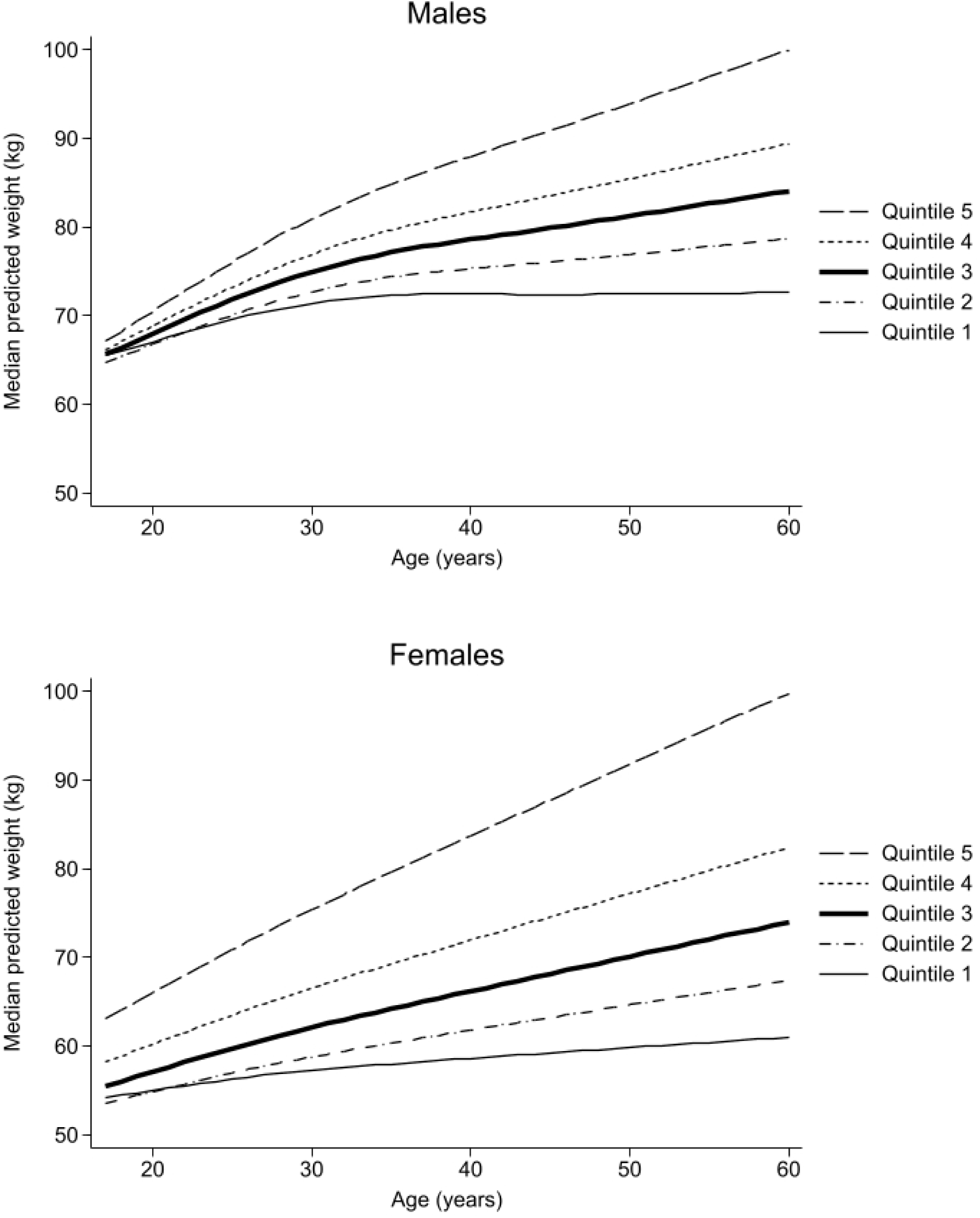
Weight Trajectories from 17 to 60 Years of Age Across Quintiles. Notes: Trajectories were estimated based on a linear mixed effects model of weight with natural cubic splines of age (4 knots) and individual random effects for slope and intercept. Other predictors included the mode of weight measurement (objective current, self-reported current, or recalled) and whether the weight measurement was obtained from the National Medical Birth Register. In the figure, trajectories are plotted for median age-specific predicted weights across the sex-specific trajectory quintiles, which are defined based on the estimated random slopes. To ensure comparability across quintiles, all predictions used for the figure assumed that weight observations were objective current, and not from the National Medical Birth Register.

**eFigure 3.**
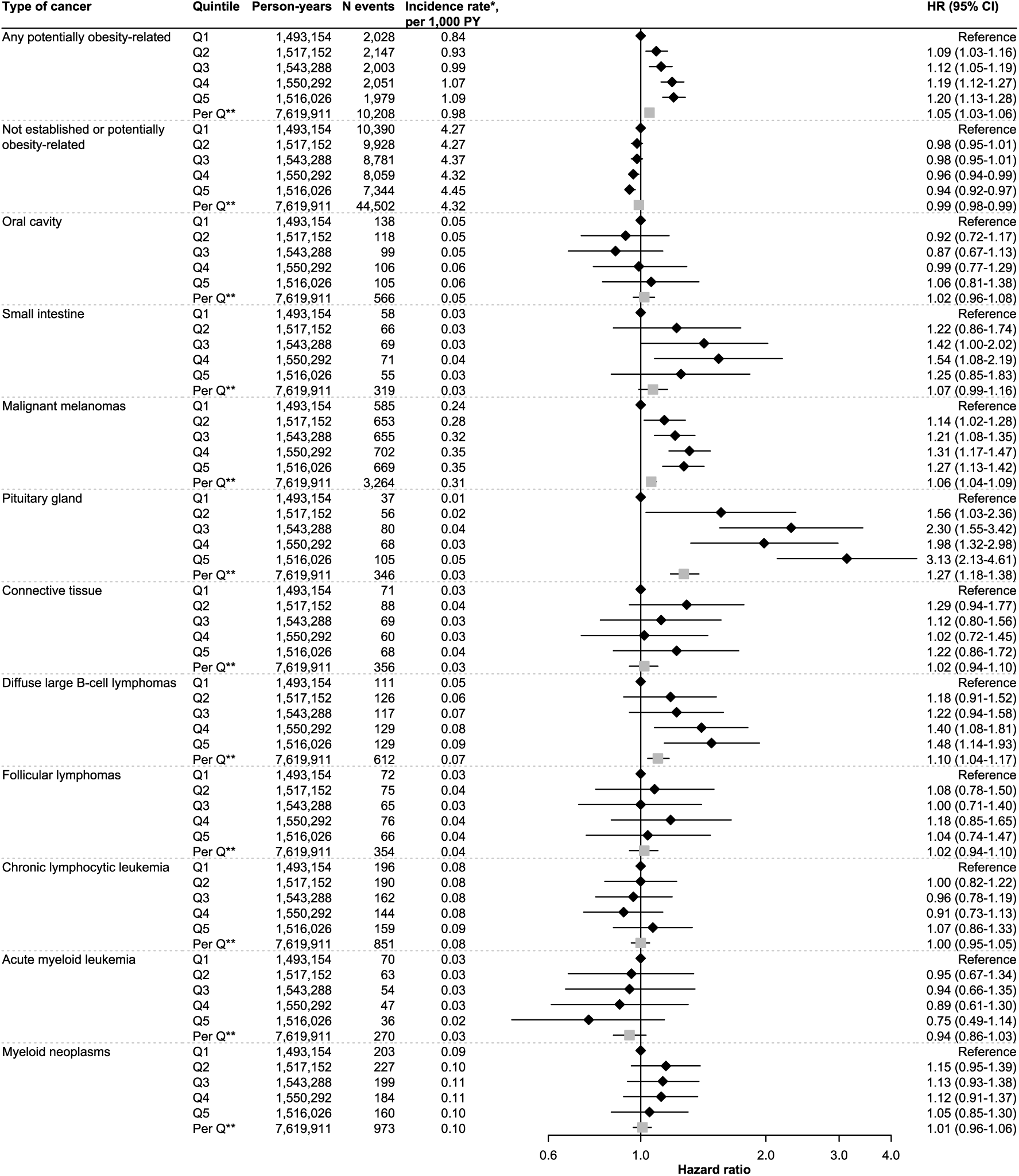
Potentially and Non-Obesity-Related Cancers, by Weight Trajectory Quintile, Males. Hazard ratios (HRs) with 95% confidence intervals (CIs) for any and different sites of potentially obesity-related cancers as well as any not established or potentially obesity-related cancer, by quintiles (Qs) of weight trajectories between ages 17 and 60. Models were estimated with Cox regression using age as the timescale, stratification with respect to birth cohort (<1940, 1940-1949, 1950-1959, 1960-1969, 1970-1979, and ≥1980), and covariate adjustment for predicted weight at age 17, height, country of birth of the individual and their parents, smoking, marital status, and the highest attained level of education. * standardized to the age distribution of the full Swedish population in 2020; ** effect of a one-quintile increase estimated from a linear model

**eFigure 4.**
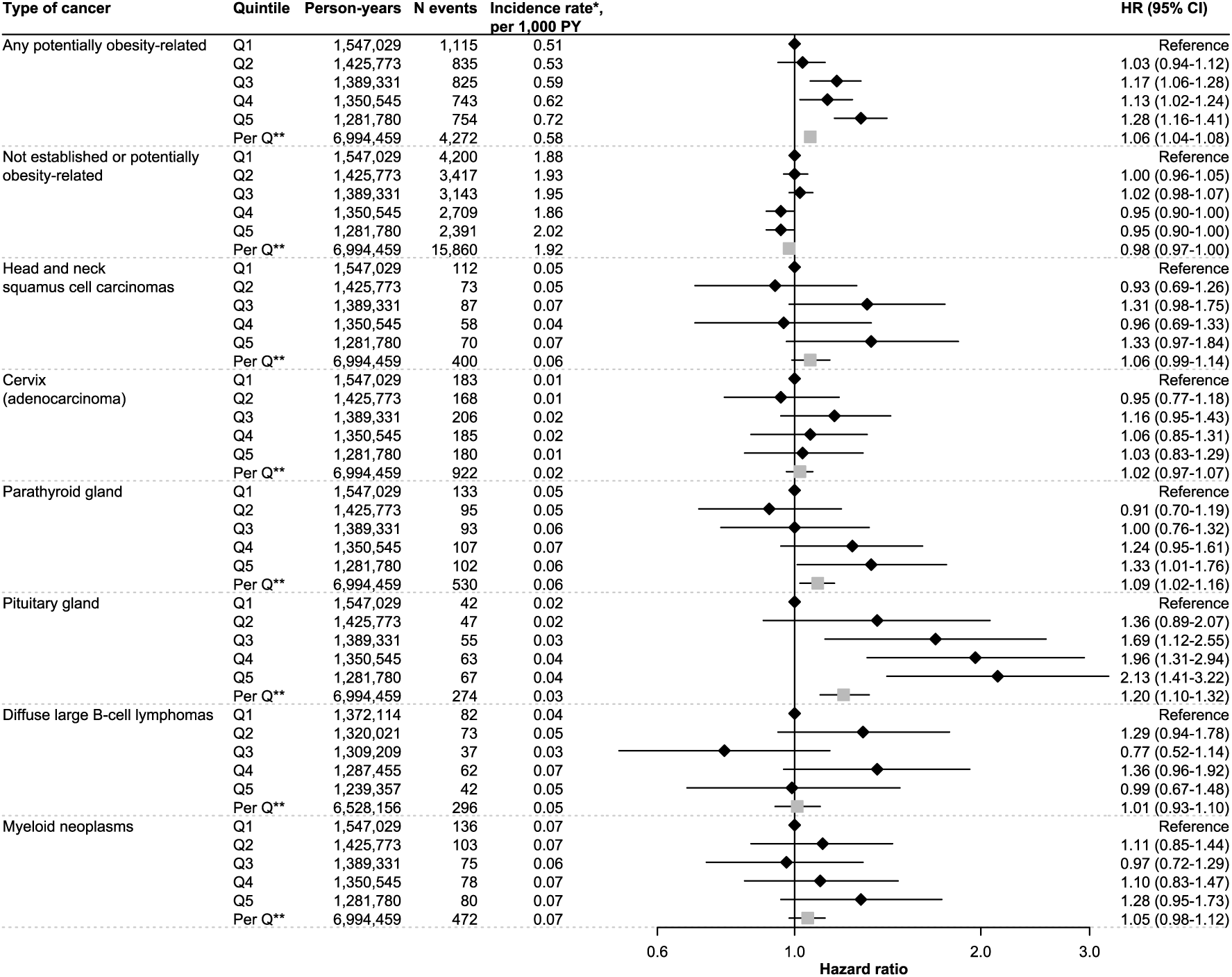
Potentially and Non-Obesity-Related Cancers, by Weight Trajectory Quintile, Females. Hazard ratios (HRs) with 95% confidence intervals (CIs) for any and different sites of potentially obesity-related cancers as well as any not established or potentially obesity-related cancer, by quintiles (Qs) of weight trajectories between ages 17 and 60. Models were estimated with Cox regression using age as the timescale, stratification with respect to birth cohort (<1940, 1940-1949, 1950-1959, 1960-1969, 1970-1979, and ≥1980), and covariate adjustment for predicted weight at age 17, height, country of birth of the individual and their parents, smoking, marital status, and the highest attained level of education. * standardized to the age distribution of the full Swedish population in 2020; ** effect of a one-quintile increase estimated from a linear model

**eFigure 5.**
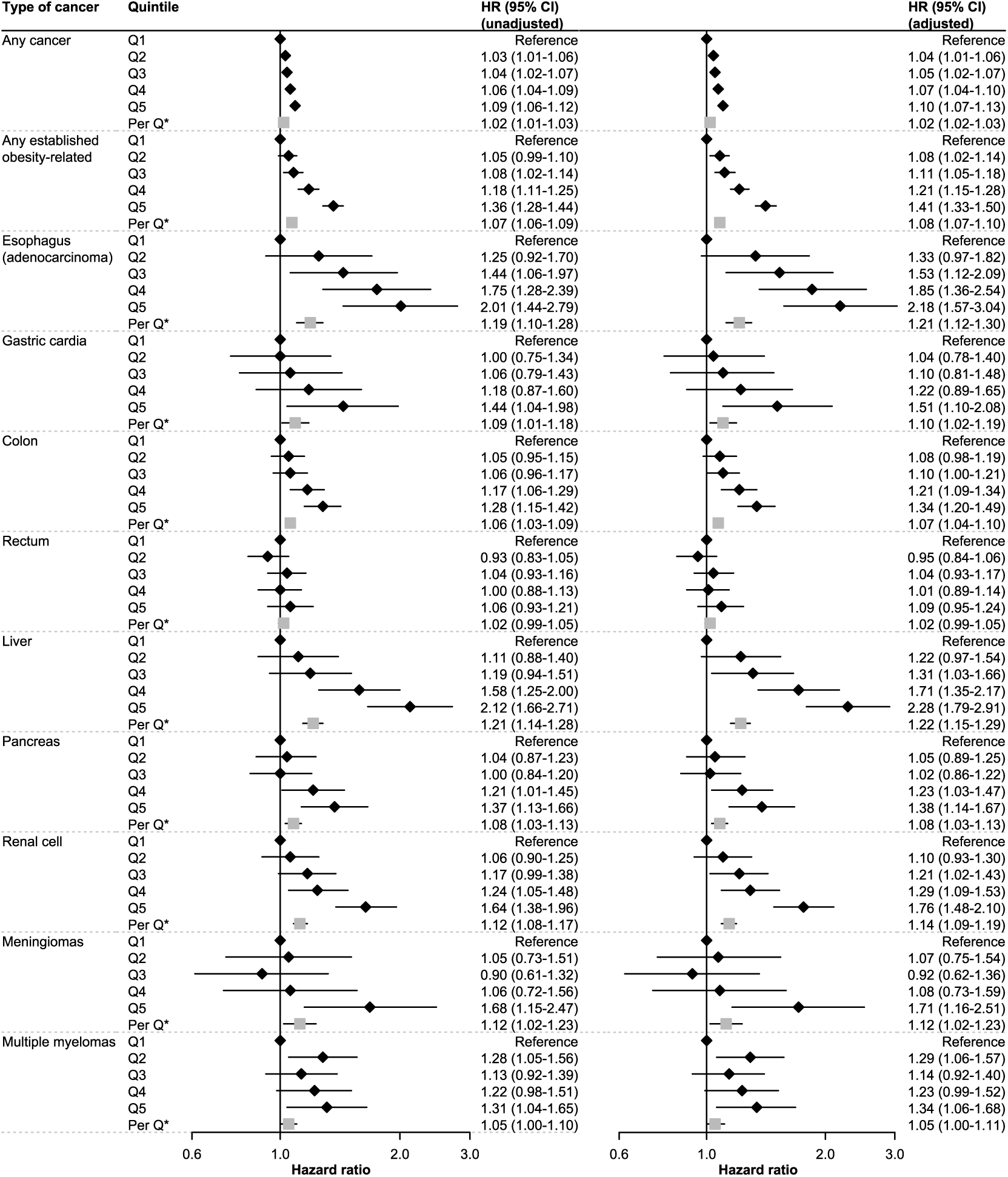
Any and Established Obesity-Related Cancers, by Age 17 Weight Quintile, Males. Hazard ratios (HRs) with 95% confidence intervals (CIs) for any cancer as well as any and different sites of established obesity-related cancers, by quintiles (Qs) of fitted weight at age 17. Results are presented with and without adjustment for subsequent quintile of weight trajectory between age 17 and 60. All models were estimated with Cox regression using age as the timescale, stratification with respect to birth cohort (<1940, 1940-1949, 1950-1959, 1960-1969, 1970-1979, and ≥1980), and covariate adjustment for predicted weight at age 17, height, country of birth of the individual and their parents, smoking, marital status, the highest attained level of education, and (in the right column) quintiles of weight trajectories. * effect of a one-quintile increase estimated from a linear model

**eFigure 6.**
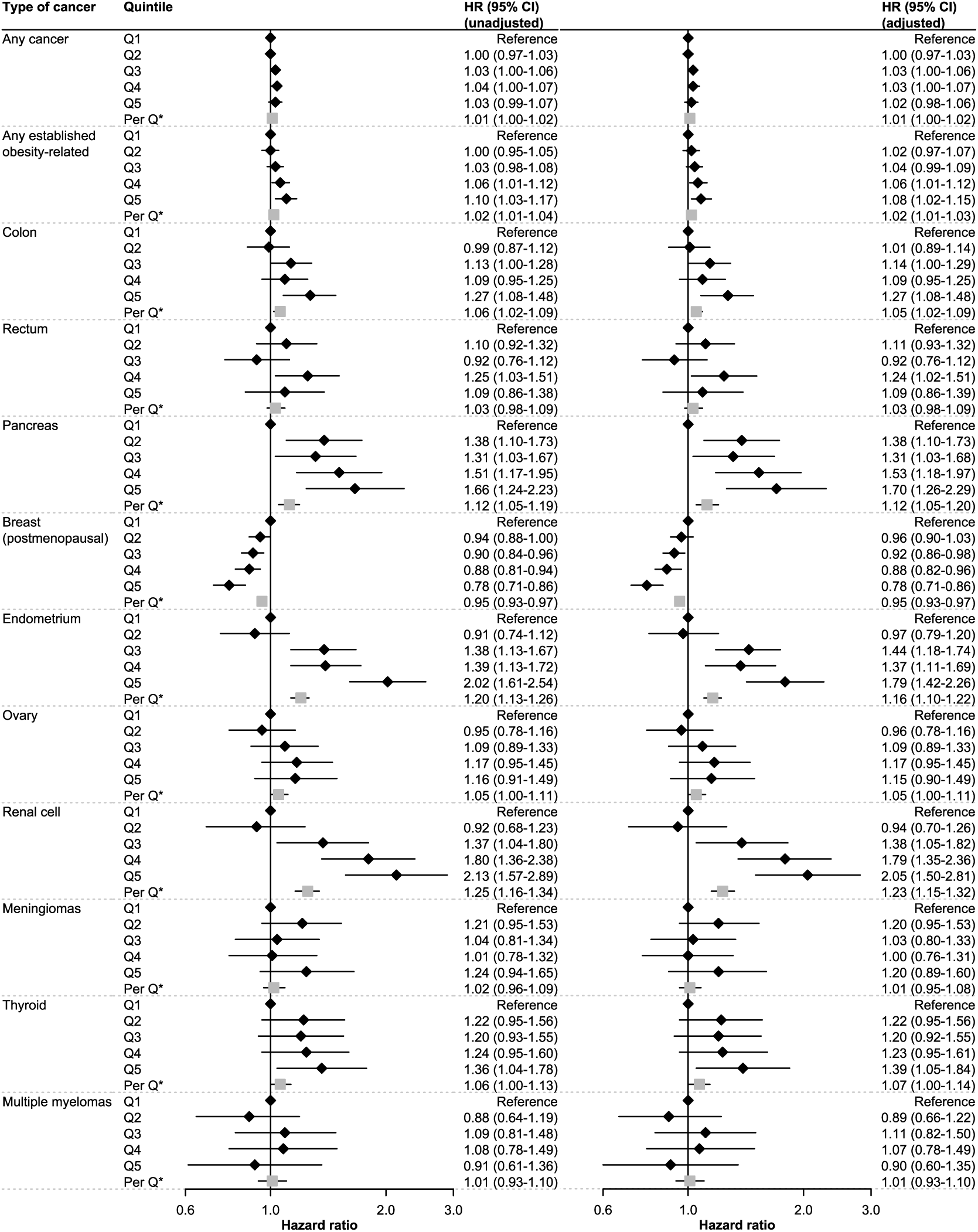
Any and Established Obesity-Related Cancers, by Age 17 Weight Quintile, Females. Hazard ratios (HRs) with 95% confidence intervals (CIs) for any cancer as well as any and different sites of established obesity-related cancers, by quintiles (Qs) of fitted weight at age 17. Results are presented with and without adjustment for subsequent quintile of weight trajectory between age 17 and 60. All models were estimated with Cox regression using age as the timescale, stratification with respect to birth cohort (<1940, 1940-1949, 1950-1959, 1960-1969, 1970-1979, and ≥1980), and covariate adjustment for height, country of birth of the individual and their parents, smoking, marital status, the highest attained level of education, and (in the right column) quintiles of weight trajectories. * effect of a one-quintile increase estimated from a linear model

**eFigure 7.**
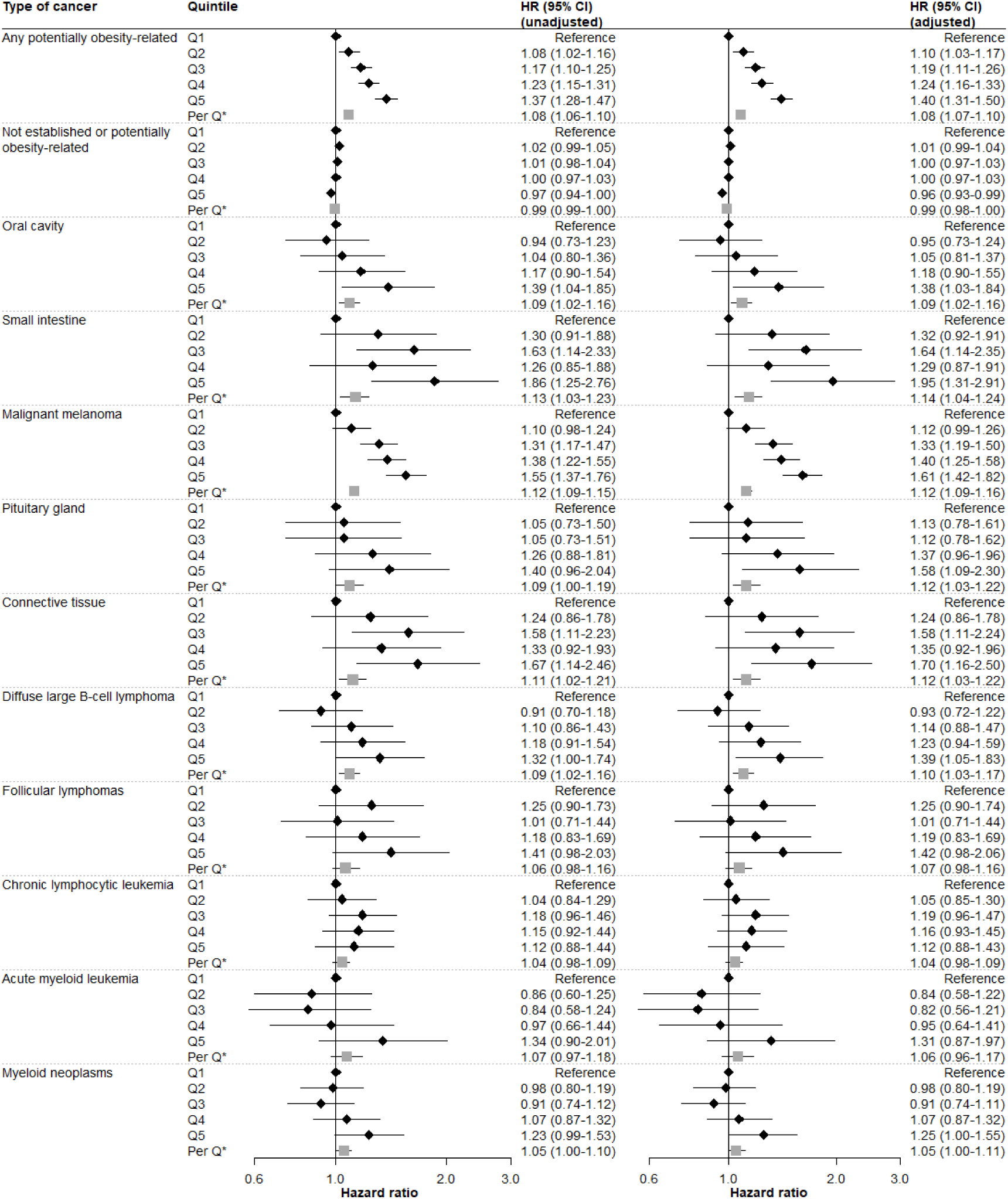
Potentially and Non-Obesity-Related Cancers, by Age 17 Weight Quintile, Males. Hazard ratios (HRs) with 95% confidence intervals (CIs) for any and different sites of potentially obesity-related cancers as well as any not established or potentially obesity-related cancer, by quintiles (Qs) of fitted weight at age 17. Results are presented with and without adjustment for subsequent quintile of weight trajectory between age 17 and 60. All models were estimated with Cox regression using age as the timescale, stratification with respect to birth cohort (<1940, 1940-1949, 1950-1959, 1960-1969, 1970-1979, and ≥1980), and covariate adjustment for height, country of birth of the individual and their parents, smoking, marital status, the highest attained level of education, and (in the right column) quintiles of weight trajectories. * effect of a one-quintile increase estimated from a linear model

**eFigure 8.**
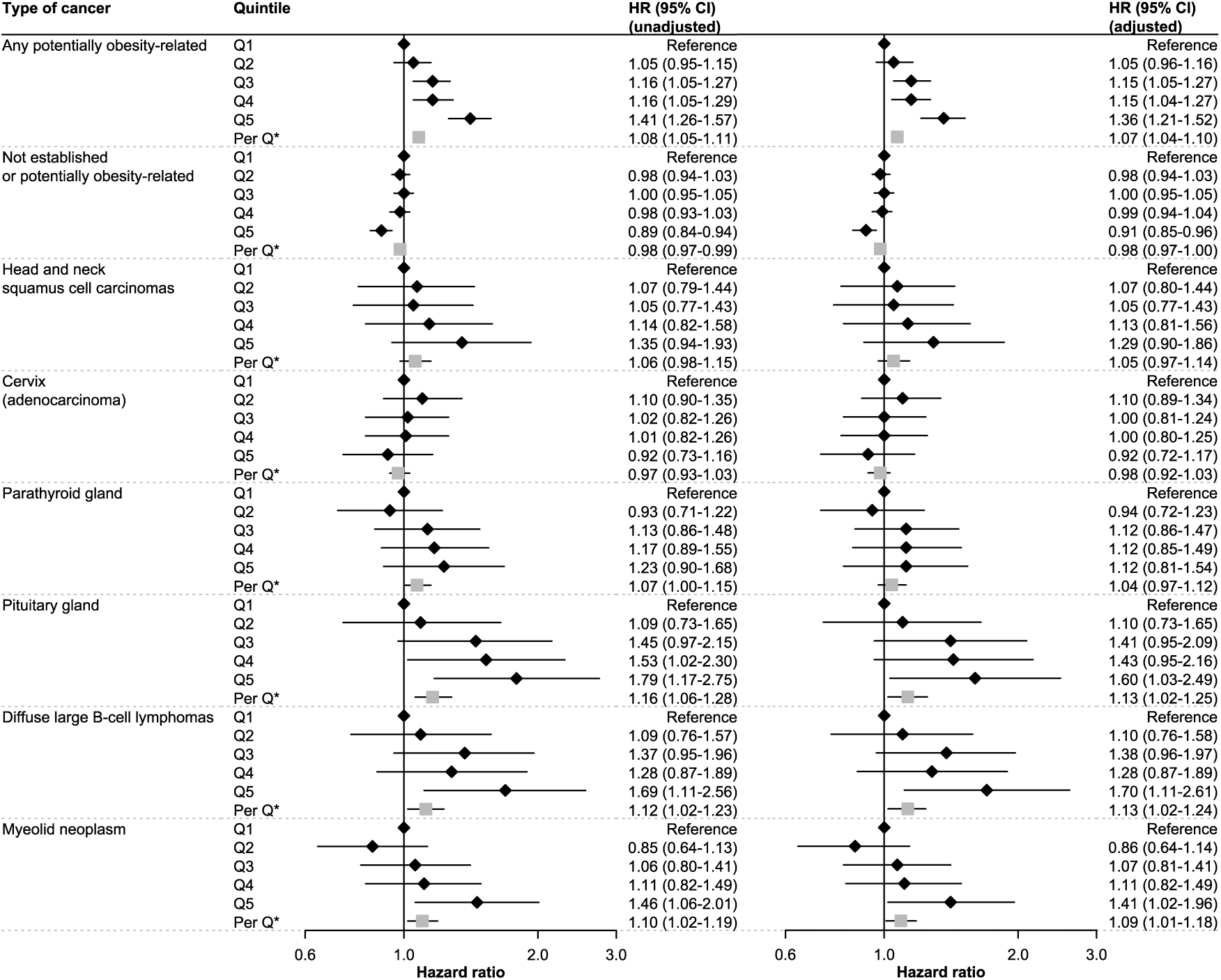
Potentially and Non-Obesity-Related Cancers, by Age 17 Weight Quintile, Females. Hazard ratios (HRs) with 95% confidence intervals (CIs) for any and different sites of potentially obesity-related cancers as well as any not established or potentially obesity-related cancer, by quintiles (Qs) of fitted weight at age 17. Results are presented with and without adjustment for subsequent quintile of weight trajectory between age 17 and 60. All models were estimated with Cox regression using age as the timescale, stratification with respect to birth cohort (<1940, 1940-1949, 1950-1959, 1960-1969, 1970-1979, and ≥1980), and covariate adjustment for height, country of birth of the individual and their parents, smoking, marital status, the highest attained level of education, and (in the right column) quintiles of weight trajectories. * effect of a one-quintile increase estimated from a linear model

**eFigure 9.**
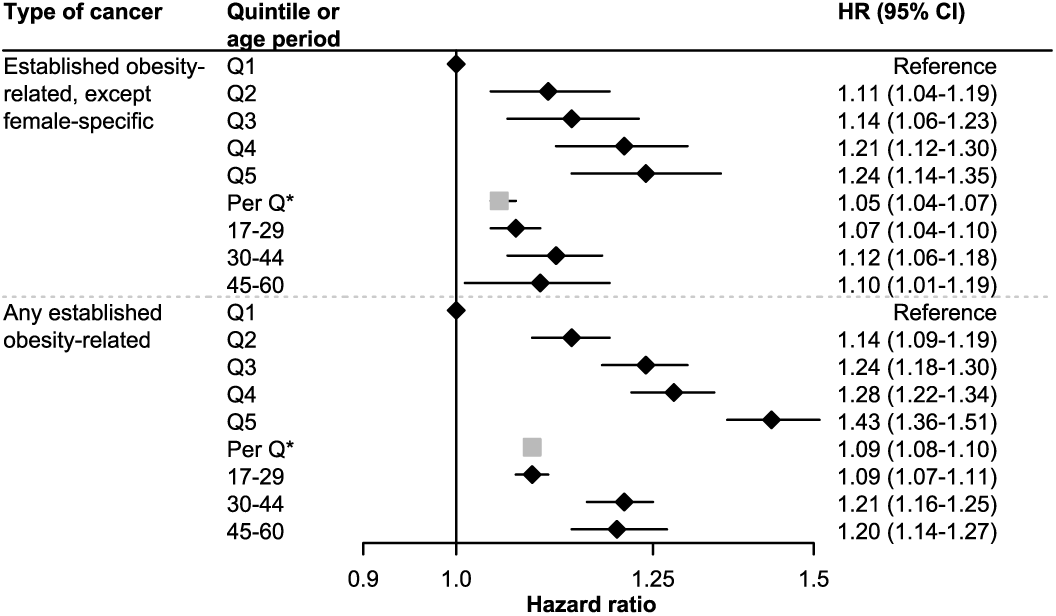
Established Obesity-Related Cancers Excluding Breast, Ovarian, and Endometrial Cancers, Females. Results are presented both for HRs across quintiles of weight change in ages 17-60 and for age interval-specific weight increases of 0.5 kg per year. HR = hazard ratio; CI = confidence interval; Q = quintile Models were estimated with Cox regression using age as the timescale, stratification with respect to birth cohort (<1940, 1940-1949, 1950-1959, 1960-1969, 1970-1979, and ≥1980), and covariate adjustment for predicted weight at age 17, height, country of birth of the individual and their parents, smoking, marital status, the highest attained level of education, and slopes of weight change in all previous age intervals. * effect of a one-quintile increase estimated from a linear model Results are contrasted with the results for all established obesity-related cancers, from the main analysis.

**eFigure 10.**
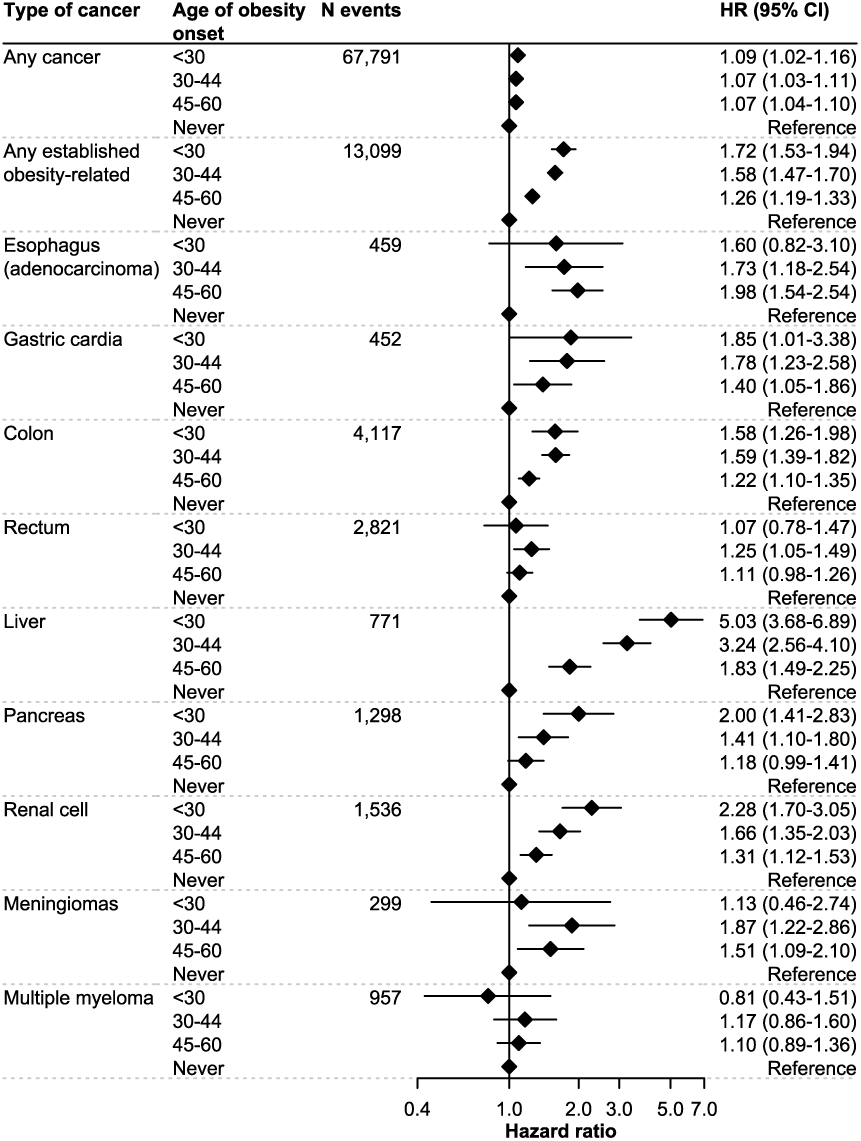
Any and Established Obesity-Related Cancers, by Age of Obesity Onset, Males. The individual’s age of obesity onset was predicted from the linear mixed effects model. Models were estimated with Cox regression using age as the timescale, stratification with respect to birth cohort (<1940, 1940-1949, 1950-1959, 1960-1969, 1970-1979, and ≥1980), and covariate adjustment for height, country of birth of the individual and their parents, smoking, marital status, and the highest attained level of education. HR = hazard ratio; CI = confidence interval

**eFigure 11.**
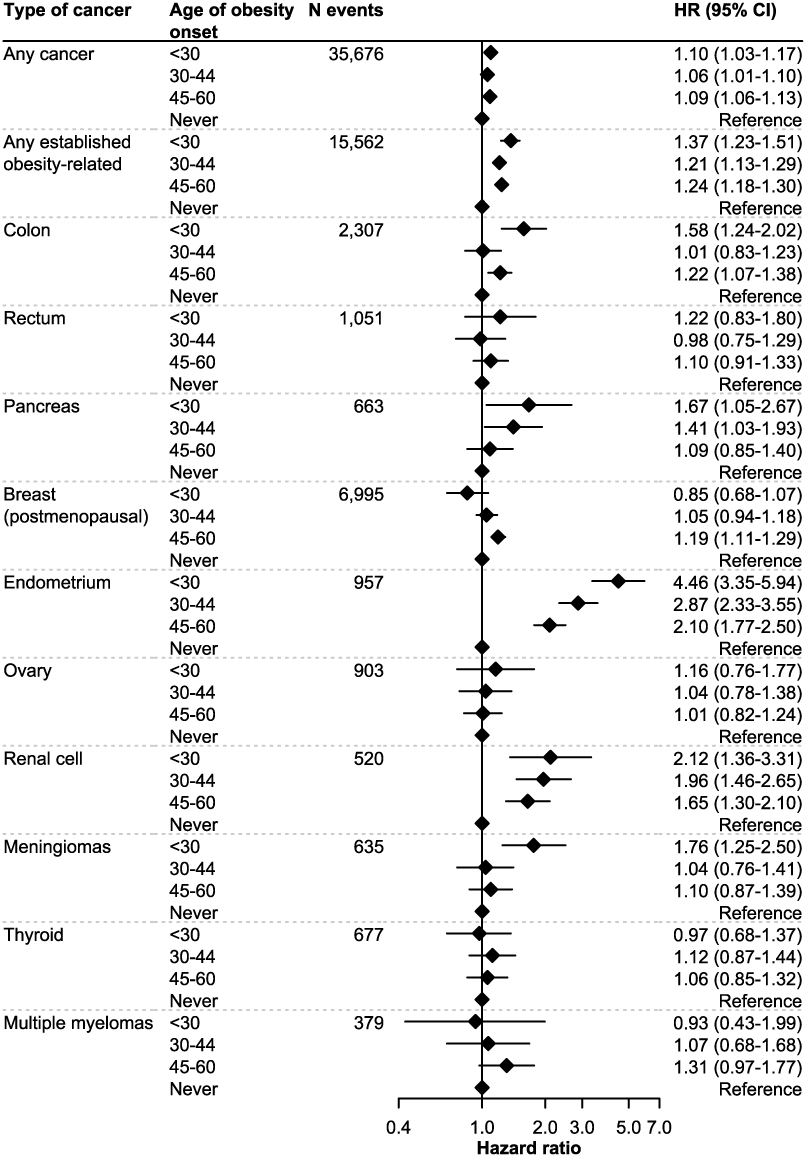
Any and Established Obesity-Related Cancers, by Age of Obesity Onset, Females. The individual’s age of obesity onset was predicted from the linear mixed effects model. Models were estimated with Cox regression using age as the timescale, stratification with respect to birth cohort (<1940, 1940-1949, 1950-1959, 1960-1969, 1970-1979, and ≥1980), and covariate adjustment for height, country of birth of the individual and their parents, smoking, marital status, and the highest attained level of education. HR = hazard ratio; CI = confidence interval

**eFigure 12.**
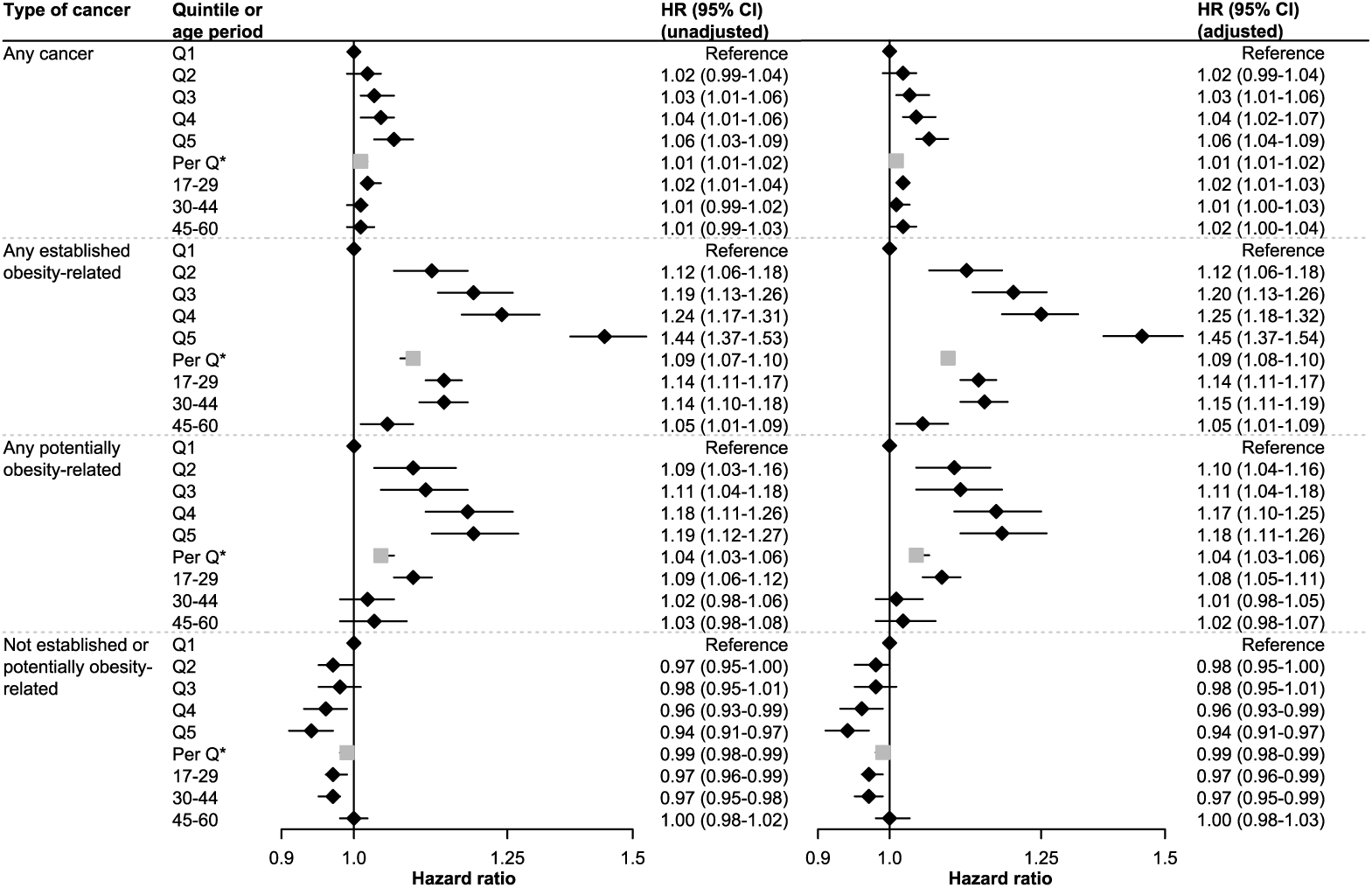
Hazard Ratios Unadjusted and Adjusted for Smoking, Males. The sample is restricted to individuals with non-missing data on smoking (98% of all males). Results are presented both for HRs across quintiles of weight change in ages 17-60 and for age interval-specific weight increases of 0.5 kg per year. HR = hazard ratio; CI = confidence interval; Q = quintile All models were estimated with Cox regression using age as the timescale, stratification with respect to birth cohort (<1940, 1940-1949, 1950-1959, 1960-1969, 1970-1979, and ≥1980), and covariate adjustment for predicted weight at age 17, height, country of birth of the individual and their parents, marital status, the highest attained level of education, slopes of weight change in all previous age intervals, and (in models to the right) smoking. * effect of a one-quintile increase estimated from a linear model

**eFigure 13.**
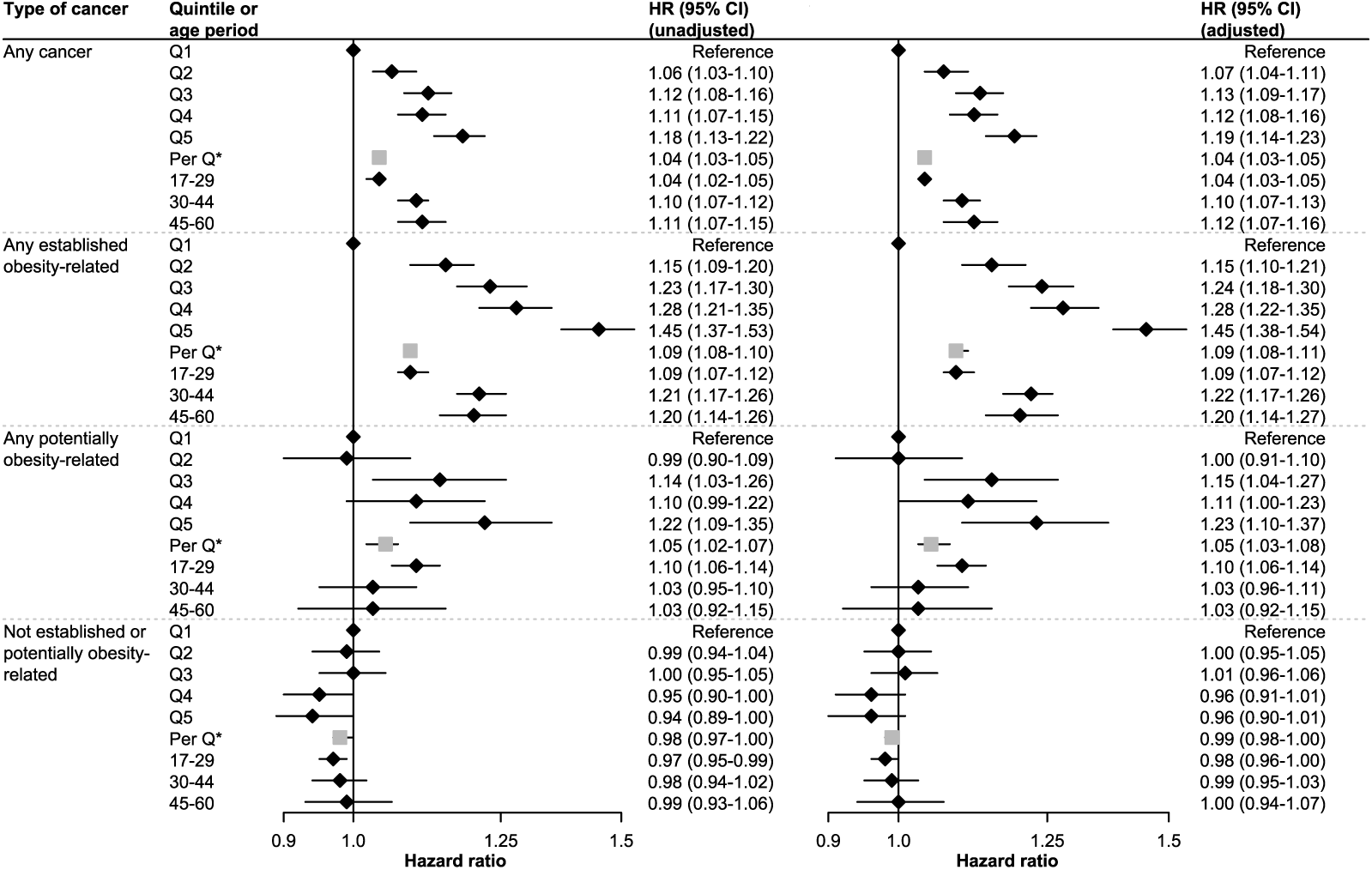
Hazard Ratios Unadjusted and Adjusted for Smoking, Females. The sample is restricted to individuals with non-missing data on smoking (85% of all females). Results are presented both for HRs across quintiles of weight change in ages 17-60 and for age interval-specific weight increases of 0.5 kg per year. HR = hazard ratio; CI = confidence interval; Q = quintile All models were estimated with Cox regression using age as the timescale, stratification with respect to birth cohort (<1940, 1940-1949, 1950-1959, 1960-1969, 1970-1979, and ≥1980), and covariate adjustment for predicted weight at age 17, height, country of birth of the individual and their parents, marital status, the highest attained level of education, slopes of weight change in all previous age intervals, and (in models to the right) smoking. * effect of a one-quintile increase estimated from a linear model

**eFigure 14.**
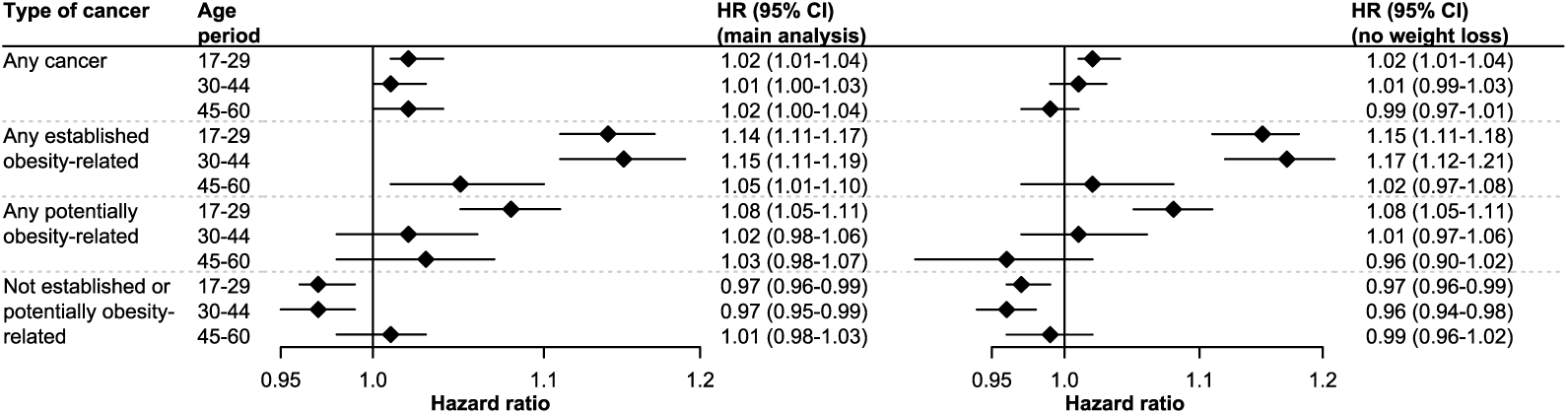
Hazard Ratios in Individuals Not Experiencing Weight Loss in Age Intervals, by Age-Specific Weight Change in Males. HRs per 0.5 kg weight increase per year. Results excluding individuals losing weight in the corresponding age interval (right) are contrasted with main analysis results (left), where individuals with weight loss were not excluded. Models were estimated with Cox regression using age as the timescale, stratification with respect to birth cohort (<1940, 1940-1949, 1950-1959, 1960-1969, 1970-1979, and ≥1980), and covariate adjustment for predicted weight at age 17, height, country of birth of the individual and their parents, marital status, the highest attained level of education, and slopes of weight change in all previous age intervals. HR = hazard ratio; CI = confidence interval The shares not experiencing weight loss were 99% (age period 17-29), 94% (age period 30-44), and 92% (age period 45-60).

**eFigure 15.**
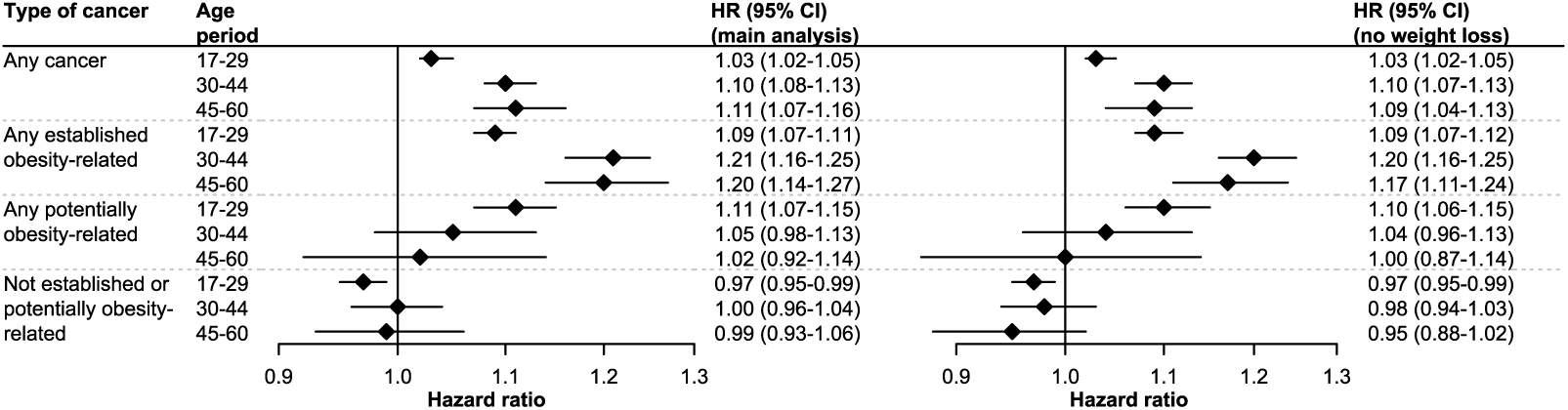
Hazard Ratios in Individuals Not Experiencing Weight Loss in Age Interval, by Age-Specific Weight Change in Females. HRs per 0.5 kg weight increase per year. Results excluding individuals losing weight in the corresponding age interval (right) are contrasted with main analysis results (left), where individuals with weight loss were not excluded. Models were estimated with Cox regression using age as the timescale, stratification with respect to birth cohort (<1940, 1940-1949, 1950-1959, 1960-1969, 1970-1979, and ≥1980), and covariate adjustment for predicted weight at age 17, height, country of birth of the individual and their parents, marital status, the highest attained level of education, and slopes of weight change in all previous age intervals. HR = hazard ratio; CI = confidence interval The shares not experiencing weight loss are 92% (age period 17-29), 97% (age period 30-44), and 98% (age period 45-60).

**eFigure 16.**
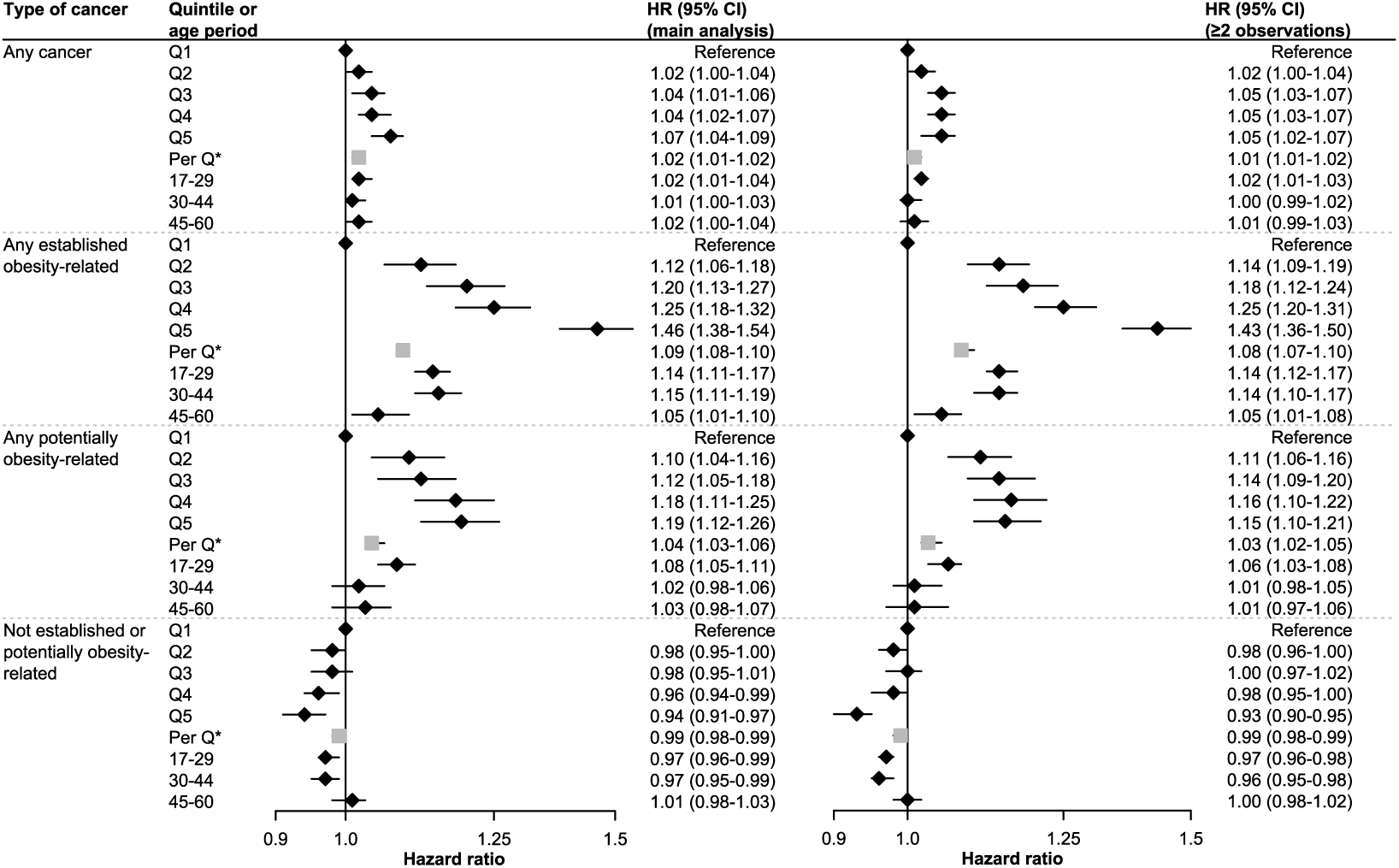
Hazard Ratios in Individuals with Two or More Weight Measurements, Males. Results are presented both for HRs across quintiles of weight change in ages 17-60 and for age interval-specific weight increases of 0.5 kg per year. Results are contrasted with results from the main analysis. Individuals in the main analyses were followed from one year after their third weight assessment, whereas individuals in the sample with at least two measurements were followed from one year after their second. Models were estimated with Cox regression using age as the timescale, stratification with respect to birth cohort (<1940, 1940-1949, 1950-1959, 1960-1969, 1970-1979, and ≥1980), and covariate adjustment for predicted weight at age 17, height, country of birth of the individual and their parents, marital status, the highest attained level of education, and slopes of weight change in all previous age intervals. The main sample of males with at least three weight assessments contains 251,041 individuals; the sample of males with at least two weight assessments contains 403,236 individuals. HR = hazard ratio; CI = confidence interval; Q = quintile * effect of a one-quintile increase estimated from a linear model

**eFigure 17.**
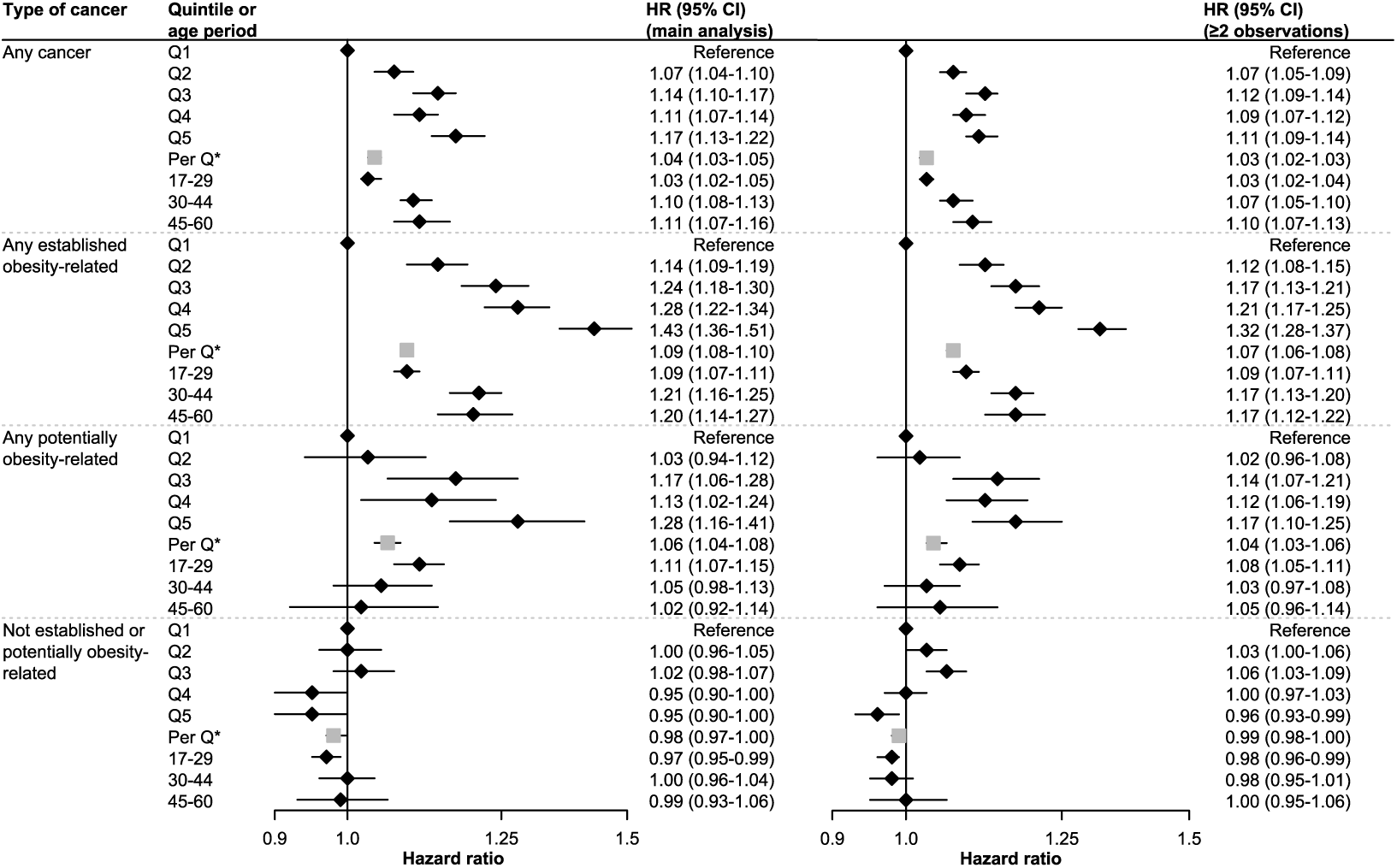
Hazard Ratios in Individuals with Two or More Weight Measurements, Females. Results are presented both for HRs across quintiles of weight change in ages 17-60 and for age interval-specific weight increases of 0.5 kg per year. Results are contrasted with results from the main analysis. Individuals in the main analyses were followed from one year after their third weight assessment, whereas individuals in the sample with at least two measurements were followed from one year after their second. Models were estimated with Cox regression using age as the timescale, stratification with respect to birth cohort (<1940, 1940-1949, 1950-1959, 1960-1969, 1970-1979, and ≥1980), and covariate adjustment for predicted weight at age 17, height, country of birth of the individual and their parents, marital status, the highest attained level of education, and slopes of weight change in all previous age intervals. The main sample of females with at least three weight assessments contains 378,981 individuals; the sample of females with at least two weight assessments contains 1,127,821 individuals. HR = hazard ratio; CI = confidence interval; Q = quintile * effect of a one-quintile increase estimated from a linear model

**eFigure 18.**
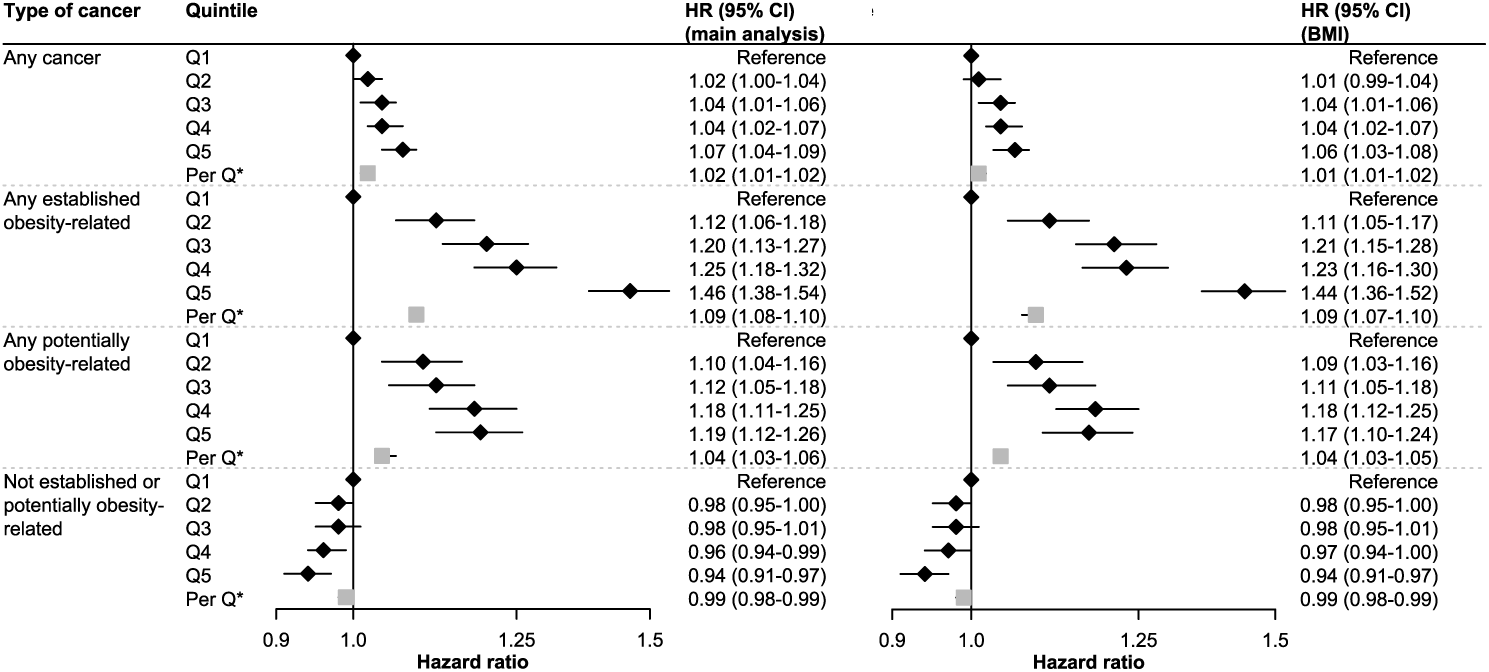
Hazard Ratios from Analyses Using BMI Trajectories, by Weight Trajectory Quintile in Males. Results for HRs across BMI (kg/m^2^) trajectories are contrasted with the results for HRs contrasting weight (kg) trajectories from the main analysis. Models were estimated with Cox regression using age as the timescale, stratification with respect to birth cohort (<1940, 1940-1949, 1950-1959, 1960-1969, 1970-1979, and ≥1980), and covariate adjustment for predicted weight at age 17, height, country of birth of the individual and their parents, marital status, and the highest attained level of education. HR = hazard ratio; CI = confidence interval; Q = quintile * effect of a one-quintile increase estimated from a linear model

**eFigure 19.**
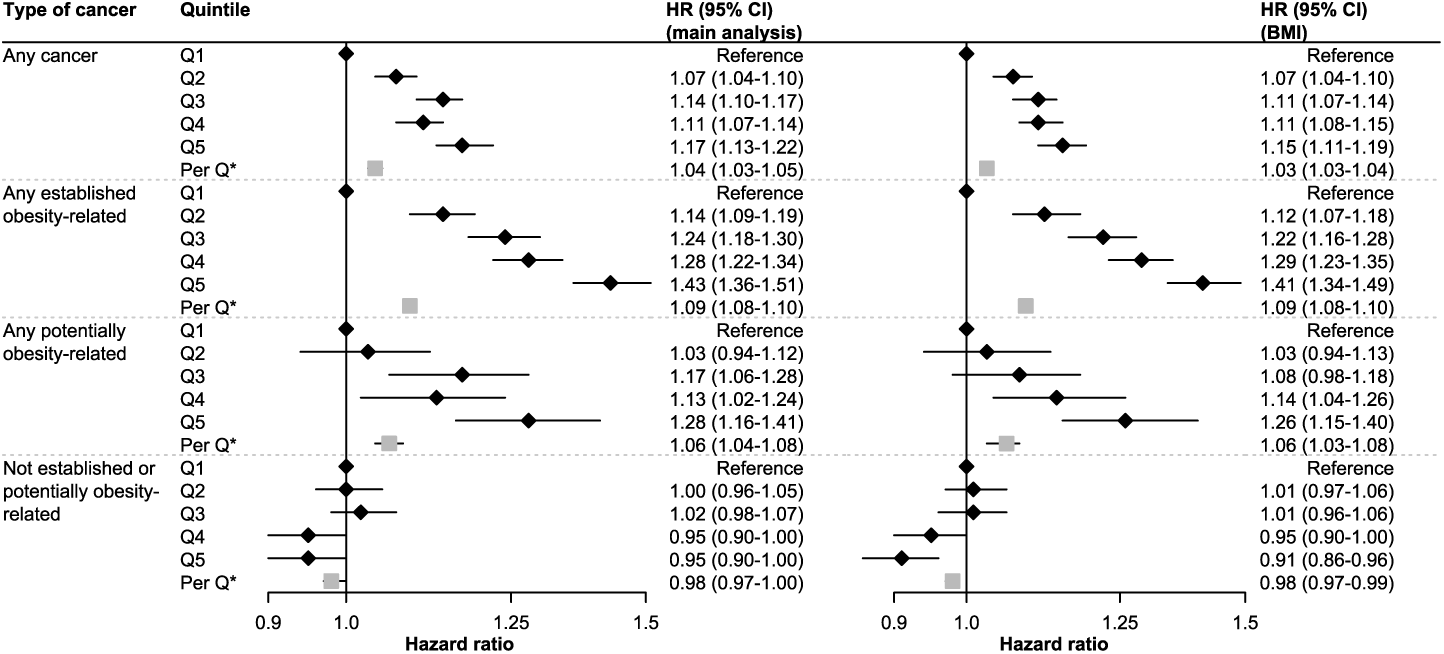
Hazard Ratios from Analyses Using BMI Trajectories, by Weight Trajectory Quintile in Females. Results for HRs across BMI (kg/m^2^) trajectories are contrasted with the results for HRs contrasting weight (kg) trajectories from the main analysis. Models were estimated with Cox regression using age as the timescale, stratification with respect to birth cohort (<1940, 1940-1949, 1950-1959, 1960-1969, 1970-1979, and ≥1980), and covariate adjustment for predicted weight at age 17, height, country of birth of the individual and their parents, marital status, and the highest attained level of education. HR = hazard ratio; CI = confidence interval; Q = quintile * effect of a one-quintile increase estimated from a linear model

